# Effect of brewers’ yeast or beta-glucan derived from *Saccharomyces cerevisiae* on breast milk supply following preterm birth: The BLOOM randomised controlled trial

**DOI:** 10.64898/2026.07.07.26357452

**Authors:** Luke E. Grzeskowiak, Lauren Williams, Alice R. Rumbold, Bronni Simpson, Renee L. Kam, Lisa N Yelland, Kathryn Dansie, Wendy V. Ingman, Amy Keir, Kathryn A. Martinello, Lisa H. Amir

## Abstract

**Objective:** Breast milk is the optimal source of nutrition for preterm infants, however, low breast milk production is common following a preterm birth. This study aimed to determine if taking brewers’ yeast or beta-glucan improves daily expressed breast milk volume.

**Design:** Randomised, blinded, parallel, placebo-controlled trial.

**Setting:** Three Australian tertiary level neonatal units.

**Patients:** Mothers with a singleton or twin pregnancy who gave birth at less than 34 weeks’ gestation.

**Interventions:** Mothers were randomised within 72 hours of birth into three parallel groups in 1:1:1 ratio to receive either brewers’ yeast, beta-glucan or placebo capsules for seven days.

**Main outcome measure:** Total expressed breast milk volume over a 24-hour period on day seven of intervention.

**Results:** A total of 105 mothers underwent randomisation between August 2022 and April 2024 (36 brewers’ yeast, 35 beta-glucan, and 34 placebo). The adjusted mean difference in daily expressed breast milk volume was 94 mL/day (95% CI -51 to 239 mL/day) between the brewers’ yeast and placebo groups, and -25 mL/day (95% CI -173 to 123 mL/day) between the beta-glucan and placebo groups. Maternal side effects were similar across groups.

**Conclusion:** We found no clear effect of short-term administration of brewers’ yeast or beta-glucan on breast-milk production following preterm birth; both interventions were well tolerated. Given the small sample size, these findings do not rule out the possibility of a clinically meaningful benefit of brewers’ yeast and suggest further research with a larger sample size may be warranted to clarify the potential clinical impact.

**Trial registration number:** ACTRN12622000968774.

**Key messages:** *What is already known on this topic:*

- Foods rich in beta-glucan, which includes brewers’ yeast derived from *Saccharomyces cerevisiae* and used in beer-making, are widely promoted based on anecdotal reports to increase breast milk production
- No randomised controlled trials have evaluated the efficacy or safety of brewers’ yeast or beta-glucan supplements for increasing breast milk production following preterm birth.

*What this study adds:*

- This randomised trial provides the first controlled data on brewers’ yeast and beta-glucan derived from *Saccharomyces cerevisiae* in comparison to placebo.
- Evidence supporting the efficacy of routine brewers’ yeast administration on improving breast milk production following preterm birth remains inconclusive, with a clinically meaningful impact unable to be ruled out. In contrast, the findings suggest beta-glucan alone appears to be unlikely to produce a meaningful difference in breast milk production.

*How this study might affect research, practice or policy:*

- Findings support the need for larger trials, with a potentially longer intervention period, to provide definitive evidence regarding the efficacy and safety of using brewers’ yeast as a galactagogue and reinforce the importance of rigorous evaluation before advocating clinical use.

## INTRODUCTION

Breast milk is the optimal enteral nutrition for preterm infants, and significantly reduces risks of mortality and major morbidities such as necrotising enterocolitis (NEC), late-onset sepsis, chronic lung disease, retinopathy of prematurity, rehospitalisation, and neurodevelopmental impairment.[1–3] For mothers, providing breast milk also offers tangible participation in infant care during separation in the Neonatal Intensive Care Unit (NICU), promoting attachment, empowerment, confidence and overall maternal wellbeing.[4–6]

Establishing and sustaining milk supply after preterm birth can be challenging. Contributing factors include physiological immaturity of the breast, inability of the infant to feed directly, NICU-related stress, and maternal co-morbidities and pregnancy complications, all commonly associated with preterm birth.[7, 8] Up to 82% of mothers of preterm infants experience delayed secretory activation[9], an established risk factor for early cessation of breastfeeding.[10] While non-pharmacological strategies (e.g., optimising use of breast milk pumps) may help, many mothers continue to face challenges with respect to adequate breast milk production.

Interest has therefore grown in the use of galactagogues to enhance breast milk production. Surveys show that over half of breastfeeding women in Australia and the United States report using galactagogues such brewers’ yeast, or substances often containing brewers’ yeast such as lactation cookies.[11, 12] Brewers’ yeast is a common name used for types of yeast used for brewing beer, with one of the most common and well-studied individual yeast strains being *Saccharomyces cerevisiae*. Brewers’ yeast is freely accessible as a nutritional supplement and available in its inactive form (dead yeast cells without fermenting properties) as a powder or capsule/tablet. While poorly studied, brewers’ yeast has a number of purported benefits to promoting lactation, with anecdotal reports of widespread promotion to lactating women and evidence that women prefer natural products over prescription medications.[13] Brewers’ yeast is high in protein, contains high levels of B vitamins and minerals (such as chromium). Chromium is thought to improve glucose homeostasis, an important factor involved in lactation.[14, 15] Further, brewers’ yeast is a source of beta-glucan (from yeast cell walls) which is thought to modulate the innate immune system,[16] with emerging evidence that altered immune activity may influence lactation.[17]

Evidence supporting the use of brewers’ yeast (or beta-glucan) derived from *Saccharomyces cerevisiae* for improving lactation remains limited, with only one identified published study.[18] A recent New Zealand trial randomised 68 breastfeeding women with a healthy singleton term infant aged 1-7 months to receive either brewers’ yeast, grown on molasses and enriched with additional nutrients, or placebo for 4 weeks.[18] While direct effects on breast milk production were not measured, no differences in the proportion of any breastfeeding and exclusive breastfeeding were evident between groups.[18] We were unable to locate any studies evaluating the use of beta-glucan.

Given increased reports of brewers’ yeast as a galactagogue and the current absence of high quality data evaluating its efficacy and safety, the main objective of this trial was to determine whether taking brewers’ yeast or beta-glucan compared with placebo leads to an increase in daily expressed breast milk volume after seven days in mothers of a preterm infant born at less than 34 weeks’ gestation.

## METHODS

### Trial design

A multicentre, randomised, controlled, clinician, researcher and participant/family blinded trial with three parallel groups was conducted at three Australian hospitals (Women’s and Children’s Hospital and Flinders Medical Centre, South Australia and The Royal Women’s Hospital, Victoria). Participants were recruited between July 2022 and April 2024, with follow-up visits and data collection completed in July 2024. The Women’s and Children’s Health Network Human Research Ethics Committee approved the study (HREC/22/WCHN/00001). The results are reported in accordance with the Consolidated Standards of Reporting Trials (CONSORT) reporting guideline.[19] The trial was registered on 8 July 2022, with the Australian and New Zealand Clinical Trials Registry (https://www.anzctr.org.au/; ACTRN12622000968774).

### Participants

Inclusion criteria were women, aged 18 years or older, who had delivered a preterm infant born at less than 34 weeks’ gestation, were between 0 and 72 h of birth, intending to provide breast milk to their preterm infant for any desired duration, and able to give informed consent. Women with a contraindication to breastfeeding such as infection with human immunodeficiency virus (HIV), as well as those with higher order pregnancies (triplet or more) were ineligible.

### Screening and consent

Depending on trial site, potential study participants were first approached by their treating nurse or midwife, or directly by a member of the study team. A member of the study team confirmed eligibility and sought voluntary, written, informed consent.

### Randomisation and blinding

Women were randomised using the REDCap Randomisation Module to one of three groups: brewers’ yeast, beta-glucan, or placebo in a 1:1:1 ratio. The randomisation schedule, stratified by study site and using randomly permuted blocks of size 6, was prepared using ralloc.ado in Stata (version 16) by an independent statistician not involved with trial participants or data.

### Intervention

Participants were randomised to one of three treatment arms, consisting of either brewers’ yeast (*Saccharomyces cerevisiae*), beta-glucan (purified from *Saccharomyces cerevisiae*) or placebo (microcrystalline cellulose). Study medications were manufactured and supplied by Leiber GmbH (Germany). Study blinding was maintained by all participants taking an oral dose of three capsules twice daily (six capsules a day) according to the treatment schedule (online supplemental Table S1), for a total of seven days. Participants were advised to commence study medications on the day following study enrolment. The intervention period commenced from the day participants started taking their study medications. To achieve the correct dosage for each treatment and maintain blinding, participants were given two colour coded bottles and took one capsule from bottle 1 and two capsules from bottle 2 in the morning and three capsules from bottle 2 in the evening. All study medications were identical in appearance.

### Outcomes

The primary outcome was expressed breast milk volume (mL) over a 24-hour period on Day 7 post study intervention. To accurately document breast milk volumes, all women were provided with 80 mL breast milk collection containers (Medela Freezing and Storage bottles) which have 1 mL gradations. Secondary outcomes, prespecified in the published protocol[20] were classified into maternal breastfeeding outcomes, safety outcomes, and infant outcomes. Maternal breastfeeding outcomes included: total expressed breast milk volume on day 21 postpartum and type of milk feeding at infant discharge or term corrected age. Safety outcomes included maternal side effects, maternal serious adverse events, maternal mortality, and infant mortality. Infant outcomes included: total cumulative volume of enteral feeds consisting of formula or donor milk (mL); proportion of enteral feeds consisting of mothers own breast milk; proportion of infants exclusively receiving mothers own breast milk as enteral feeds; weight z-score (g), length z-score (cm) and head circumference z-score (cm) at discharge or term corrected age; weight growth velocity (g/day), length growth velocity (cm/week) and head circumference growth velocity (cm/week) birth to term equivalent; and infant length of stay (number of days).

### Sample size calculation

A sample of 99 women (33 per arm) yielded 90% power, 0.025 alpha (type I error) to show a difference in the mean daily breast milk volume of 150 mL/day (with a standard deviation of 200 mL/day) between each of the intervention arms and the control arm. The calculation accounted for 10% loss to follow-up and incorporated an adjustment for a 0.6 correlation between baseline breast milk volume and volume on Day 7.

### Data collection

Data were collected from participants across four study time-points (Baseline, Day 7 post intervention, Day 21 postpartum, and at infant discharge or at term corrected age [whichever occurred first]). Questionnaires and assessments were completed at study visits as described in the published protocol.[20] Study data were collected and managed using REDCap electronic data capture tools hosted at SAHMRI.[21, 22]

### Statistical analysis

Data analysis was performed by an independent statistician (KD), after locking the database and unblinding the treatment codes, at SAHMRI, Adelaide, Australia, using Stata version 19.5 (College Station, TX: StataCorp LP). Analysis followed the prespecified SAP (**online supplemental appendix 1**) and was performed using an intention-to-treat approach. Participants found to be ineligible after randomisation remained in the analysis population. No interim analyses were planned or performed.

The primary outcome (mean daily expressed breast milk volume measured on day 7 post intervention) and all continuous secondary outcomes were analysed using linear regression, with the effect of treatment described as a mean difference with a 95% confidence interval.

Binary secondary outcomes were analysed using log binomial regression, with the effect of treatment described as a relative risk with a 95% confidence interval; Fisher’s exact test was used instead for rare events. An unplanned analysis of infant length of stay using negative binomial regression was performed due to the highly skewed distribution of the outcome, with the effect of treatment described as an adjusted incidence rate ratio with 95% confidence interval. For infant outcomes, clustering due to multiple births was taken into account using generalised estimating equations with an independence working correlation structure.

Analyses were adjusted for the stratification variable (study site) unless convergence problems occurred or a Fisher’s exact test was performed. For the primary outcome, adjustment was additionally made for expressed breast milk volume from a single expressing episode prior to commencing study medications.

For the primary outcome, missing data were addressed using multiple imputation implemented under a missing at random (MAR) assumption as outlined in the Statistical Analysis Plan. A complete case analysis was also conducted to assess the robustness of findings. For secondary outcomes, analyses were performed on the available data only.

For the primary outcome only, analyses were performed to test for evidence of effect modification by (1) plurality (singleton vs. twins), (2) parity (primiparous vs. multiparous), (3) infant gestation at birth (23+0–29+6 vs 30+0–31+6 vs 32+0-33+6 weeks’ gestation), (4) maternal body mass index (underweight/normal weight vs overweight/obese). Effect modification was assessed by including the subgroup variable as well as its interaction with treatment group in the linear regression model. Interaction p-values are reported, along with treatment effect estimates and confidence intervals for each subgroup regardless of the statistical significance of the interaction.

A p-value of < 0.025 was considered statistically significant to account for the two planned comparisons per outcome (brewers’ yeast vs placebo and beta-glucan vs placebo).

## RESULTS

### Recruitment, characteristics of participants and study adherence

A total of 105 women were randomly assigned at three trial centres (36 to brewers’ yeast, 35 to beta-glucan, and 34 to placebo). After loss to follow-up, primary outcome data were available for 87 (83%) women, however 4 women did not have baseline breast milk volume required for adjustment in the primary analysis, so the complete case analysis was performed on 83 (79%) women. After multiple imputation for missing data, 105 women were included in the primary outcome analysis (**Figure 1**).

**Figure 1:**
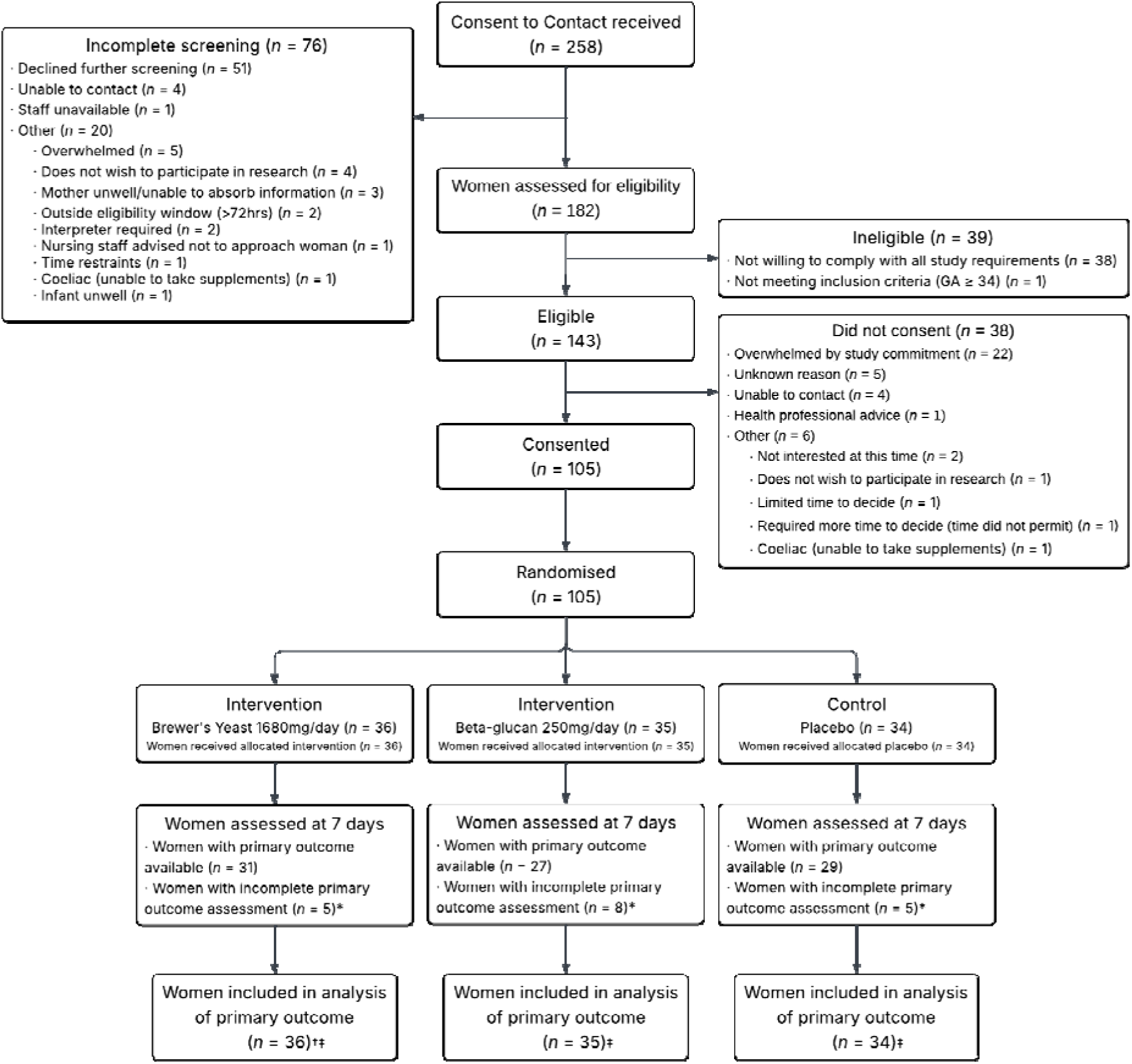
Flowchart summarising consent to contact, screening, enrolment, randomisation and analysis in the BLOOM randomised controlled trial. *Includes participants withdrawn from the study but approved the use of their data collected prior to withdrawal and hence all were included in the analysis population ^†^Includes ineligible participant randomised outside of 72 hours post birth protocol window (randomised at 83 hours post birth) ^‡^Includes multiple imputation for mothers with missing data

The mean maternal age was 32.5 (SD 5.7) years, 52 mothers (50%) had no prior births, while 12 (11%) had a multiple pregnancy. The median gestation at birth was 30.0 (IQR 28.0-32.0) weeks, while 77 (73.0%) gave birth by caesarean section. Women were recruited within a mean of 41.0 hours (SD 18.3) since birth, with a median time to first breast milk expression of 5.0 hours (IQR 3.0, 10.0) and median baseline breast milk volume from a single expression prior to randomisation of 7 mL (IQR 2-23). Baseline characteristics were reasonably balanced between the treatment groups (**Table 1**).

**Table 1:** Baseline characteristics of mothers and infants included in study by treatment group.

| Maternal characteristics | Brewers' Yeast<br>N=36 | Beta-glucan<br>N=35 | Placebo<br>N=34 | Total<br>N=105 |
| --- | --- | --- | --- | --- |
| Recruitment site |  |  |  |  |
| Women's and Children's Hospital | 21 (58) | 21 (60) | 22 (65) | 64 (61) |
| Flinders Medical Centre | 11 (31) | 10 (29) | 10 (29) | 31 (30) |
| Royal Women's Hospital | 4 (11) | 4 (11) | 2 (6) | 10 (10) |
| Age, years [mean (SD)] | 33.3 (5.6) | 32.2 (5.6) | 31.9 (5.9) | 32.5 (5.7) |
| Primiparous (birth > 20 weeks) | 14 (39) | 20 (57) | 18 (53) | 52 (50) |
| Multiple pregnancy | 5 (14) | 4 (11) | 3 (9) | 12 (11) |
| Pre-pregnancy BMI, kg/m<sup>2</sup> [median (IQR)] | 24.5 (22.4, 30.9) | 26.3 (23.2, 31.6) | 27.4 (23.7, 33.5) | 25.9 (23.1, 32.4) |
| Born in Australia | 26 (72) | 28 (80) | 21 (62) | 75 (71) |
| Aboriginal or Torres Strait Islander | 3 (8) | 1 (3) | 3 (9) | 7 (7) |
| Completed secondary education | 30 (83) | 30/34 (88) | 27 (79) | 87/104 (84) |
| Completed further study | 29 (81) | 32 (91) | 29 (85) | 90 (86) |
| Annual household income |  |  |  |  |
| ≤ \$50,000 | 5 (14) | 2 (6) | 5 (15) | 12 (11) |
| \$50,001 to \$150,000 | 4 (11) | 5 (14) | 4 (12) | 13 (12) |
| \$100,001 to \$150,000 | 9 (25) | 10 (29) | 7 (21) | 26 (25) |
| > \$150,001 | 12 (33) | 7 (20) | 11 (32) | 30 (29) |
| Undisclosed | 6 (17) | 11 (31) | 7 (21) | 24 (23) |
| Current smoker | 2 (6) | 1 (3) | 4 (12) | 7 (7) |
| Conceived using fertility treatment | 7 (19) | 4 (11) | 4 (12) | 15 (14) |
| Pregnancy Complications |  |  |  |  |
| Pre-pregnancy diabetes | 1 (3) | 0 (0) | 2 (6) | 3 (3) |
| Gestational diabetes | 12 (33) | 9 (26) | 10 (29) | 31 (30) |
| Hypertensive disorders of pregnancy | 7 (19) | 5 (14) | 7 (21) | 19 (18) |
| Intrauterine growth restriction | 7 (19) | 8 (23) | 7 (21) | 22 (21) |
| Gestation at birth, week [median (IQR)] | 31.0 (29.0, 32.0) | 30.0 (27.0, 31.0) | 31.0 (28.0, 32.0) | 30.0 (28.0, 32.0) |
| Caesarean section birth | 25 (69) | 28 (80) | 24 (76) | 77 (73) |
| Hours since birth at time of randomisation [median (IQR)] | 36.5 (25.5, 56.0) | 38.0 (24.0, 54.0) | 44.5 (28.0, 59.0) | 38.0 (27.0, 56.0) |
| Time to first breast milk expression following birth [median (IQR)] | 6.0 (3.0, 11.0) | 4.0 (1.0, 6.0) | 6.0 (3.0, 12.0) | 5.0 (3.0, 10.0) |
| Baseline breast milk volume from single expression prior to intervention, | 8.0 (3.5, 30.0) | 5.8 (2.4, 15.0) | 4.0 (2.1, 22.0) | 6.5 (2.4, 23.0) |

| **Maternal characteristics** | **Brewers' Yeast**<br>**N=36** | **Beta-glucan**<br>**N=35** | **Placebo**<br>**N=34** | **Total**<br>**N=105** |
| --- | --- | --- | --- | --- |
| mL [median (IQR)] |  |  |  |  |
| **Infant characteristics** | **Brewers' Yeast**<br>**N=41** | **Beta-glucan**<br>**N=39** | **Placebo**<br>**N=37** | **Total**<br>**N=117** |
| Male infant sex | 21 (51) | 23 (59) | 32 (57) | 65 (56) |
| Birth weight (g), mean (SD) | 1582.8 (443.7) | 1323.7 (475.4) | 1536.8 (647.3) | 1481.9 (533.8) |
| Birth length (cm), mean (SD) | 41.5 (3.85) | 38.3 (4.68) | 39.5 (6.25) | 39.8 (5.1) |
| Head circumference (cm),<br>mean (SD) | 28.7 (2.19) | 27.1 (3.05) | 28.4 (3.94) | 28.1 (3.2) |

Values are n (%) unless otherwise indicated.

During the intervention period women were expressing a median of 5 (IQR 4, 6) times each day and most were using a double breast pump. Almost 1 in 7 women reported taking a galactagogue in addition to the study intervention. Post-randomisation characteristics were reasonably balanced between the treatment groups (**Table 2**).

**Table 2:** Post-randomisation characteristics during the intervention period.

| Maternal variable | Brewers' Yeast<br>N=36 | Beta-glucan<br>N=35 | Placebo<br>N=34 | Overall<br>N=105 |
| --- | --- | --- | --- | --- |
| Average number of breast milk expression episodes per day [median (IQR)] | 5.4 (2.4, 5.8) | 5.1 (3.9, 6.0) | 5.5 (4.2, 6.3) | 5.4 (3.9, 6.1) |
| Frequency of double breast expression |  |  |  |  |
| Hardly ever (>0-20% times a day) | 1 (3) | 4 (11) | 2 (6) | 7 (7) |
| Sometimes (21-80% times a day) | 6 (17) | 3 (9) | 2 (6) | 11 (10) |
| Mostly/Always (81-100% of times a day) | 22 (61) | 17 (49) | 20 (59) | 59 (56) |
| Missing | 7 (19) | 11 (31) | 10 (29) | 28 (27) |
| Infant(s) ever put to the breast | 27 (75) | 26 (74) | 25 (74) | 77 (73) |
| Any galactagogue (prescription or non-prescription) taken | 7 (19) | 3 (9) | 6 (18) | 16 (15) |
| Type of galactagogue taken |  |  |  |  |
| Domperidone | 0 (0) | 0 (0) | 2 (6) | 2 (2) |
| Brewers' yeast | 3 (8) | 0 (0) | 1 (3) | 4 (4) |
| Lactation cookies | 3 (8) | 1 (3) | 1 (3) | 5 (5) |
| Oats | 3 (8) | 1 (3) | 1 (3) | 5 (5) |
| Lactation tea/drink | 1 (3) | 0 (0) | 2 (6) | 3 (3) |
| Ginger | 2 (6) | 0 (0) | 0 (0) | 2 (2) |
| Flaxseed | 2 (6) | 0 (0) | 1 (3) | 3 (3) |
| Type of side effects reported<sup>‡</sup> |  |  |  |  |
| Restlessness/trouble sleeping | 0 (0) | 1 (3) | 0 (0) | 1 (1) |
| Dizziness | 0 (0) | 0 (0) | 1 (3) | 1 (1) |
| Constipation | 0 (0) | 0 (0) | 1 (3) | 1 (1) |
| Diarrhoea | 2 (6) | 1 (3) | 3 (9) | 6 (6) |
| Abdominal pain/cramping | 3 (8) | 0 (0) | 1 (3) | 4 (4) |
| Other | 2 (6) | 3 (9) | 1 (3)<sup>§</sup> | 6 (6) |
| Unblinded before primary outcome assessment | 0 (0) | 0 (0) | 0 (0) | 0 (0) |
| Which study group believed to be in |  |  |  |  |
| Brewers' Yeast/Beta-glucan | 15 (42) | 4 (11) | 7 (21) | 26 (25) |
| Placebo | 1 (3) | 6 (17) | 3 (9) | 10 (10) |
| Unknown/Not sure | 12 (33) | 18 (51) | 19 (56) | 49 (47) |
| Missing | 8 (22) | 7 (20) | 5 (15) | 20 (19) |

Values are n (%) unless otherwise indicated.
‡ "Type of side effects reported" calculated as the number of times a particular symptom was reported by unique participants at any time point. Multiple reports of the same symptom by a single participant were only counted once.
§ "Type of side effects reported" as "other" occurred three times in one participant, and all three "other" side effects were different

Adherence to the intervention was modest, with 60 (57%) women reporting taking 90% or more of the allocated doses during the intervention period (**online supplemental Table S2**).

Adherence appeared to be slightly higher for those in the brewers’ yeast group (26/36 (72%)) compared with the beta-glucan group (18/35 (51%)) or placebo group (16/34 (47%)).

### Outcomes

#### Primary outcome

On day 7 post intervention, mean daily expressed breast milk volume (over a 24-hour period) was 598 mL (SD 320 mL), 430 mL (273 mL) and 474 mL (361 mL) for those assigned to brewers’ yeast, beta-glucan, or placebo, respectively. Following multiple imputation, the mean difference between brewers’ yeast and placebo was 94 mL (95% Confidence Interval (CI), -51 to 239; *P* = 0.20) and between beta-glucan and placebo was -25 mL (95% CI, -173 to 123; *P* = 0.74) (**Table 3**). These findings were consistent with the complete case analysis (**online supplemental Table S3**).

**Table 3:**
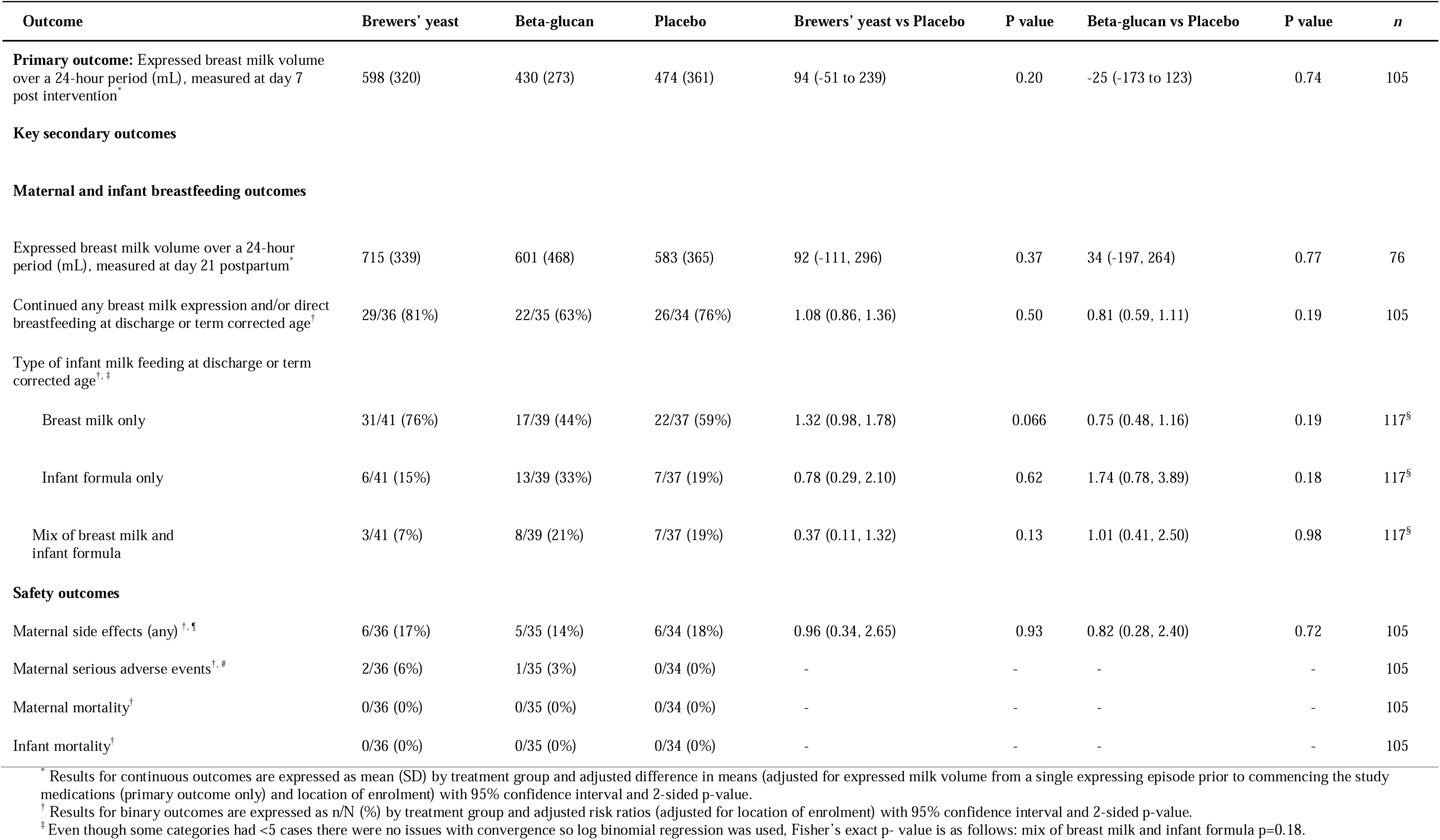
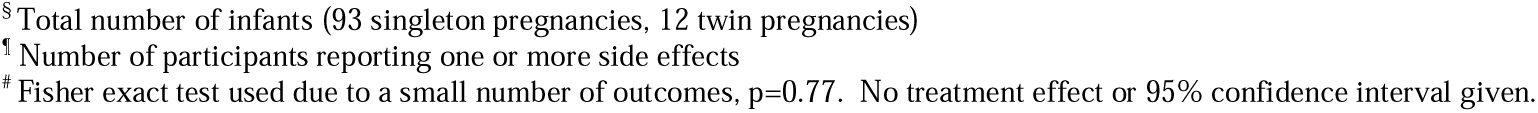
Trial outcomes.

Results of the prespecified subgroup analyses are reported in **online supplemental Table S4**, showing no clear evidence of effect modification by plurality, parity, gestation at birth, or body mass index.

### Maternal and infant breastfeeding outcomes

At day 21 postpartum, mean daily expressed breast milk volume (over a 24-hour period) was 715 mL (SD 339 mL), 601 mL (468 mL) and 583 mL (365 mL) for those assigned to brewers’ yeast, beta-glucan, or placebo, respectively (**Table 3**). The mean difference between brewers’ yeast and placebo was 92 mL (95% CI, -111 to 296 mL; *P* = 0.37) and between beta-glucan and placebo was 34 mL (95% CI, -197 to 264 mL; *P* = 0.77). The proportion of women continuing to breastfeed at infant discharge or term corrected age was similar across treatment groups (81%, 63% and 76% for brewers’ yeast, beta-glucan, and placebo, respectively (**Table 3**)). At infant discharge or term corrected age, the proportion of infants receiving breast milk only was 76% for those receiving brewers’ yeast, 44% for those receiving beta-glucan, and 59% for those receiving placebo (Brewers’ yeast vs placebo Risk Ratio (RR) 1.32; 95% CI 0.98-1.78; beta-glucan vs placebo RR 0.75; 95% CI 0.48-1.16) (**Table 3**).

### Safety outcomes

Maternal side effects were reported at a similar frequency across all three groups (**Table 2**). Three maternal serious adverse events occurred during the trial all related to hospital admissions. All were deemed to be related to postnatal complications rather than study treatment. There were no cases of maternal or infant mortality (**Table 3**).

### Infant outcomes

There were no significant between-group differences in secondary infant feeding outcomes, including cumulative volume of enteral feeds continuing formula, donor milk or mother’s own breast milk, exclusive provision of mothers’ own breast milk during infant hospitalisation, weight, length or head circumference z-scores, or length of hospital stay (**online supplemental Table S5**).

## DISCUSSION

We found inconclusive evidence that short-term administration of brewers’ yeast or beta-glucan following preterm birth improves breast milk production, while both interventions appear well tolerated. There were also no clear differences in any of the secondary maternal or infant outcomes assessed as part of the trial.

To our knowledge, this is the first RCT to evaluate the effect of brewers’ yeast on breast milk production following preterm birth. A recent trial randomised 68 breastfeeding women with a healthy singleton term infant aged 1-7 months to receive brewers’ yeast or placebo for 4 weeks.[18] While a greater proportion of women receiving brewers’ yeast reported a perceived increase in breast milk production compared to those receiving placebo, direct effects on breast milk production were not measured and no differences in the proportion of any breastfeeding and exclusive breastfeeding were evident between groups.[18] Therefore the impacts of *Saccharomyces cerevisiae* yeast-based supplements on breast milk production remain unclear and may differ according to substrate type, production methods, dose, and duration of treatment. Currently there are no standards or regulations regarding the names, sources, or dosage of brewers’ yeast containing products.

The brewers’ yeast used in this trial was *Saccharomyces cerevisiae* and was sourced as a by-product from brewers’ fermenting cellars, with the beta-glucan isolated from the same yeast product.[23] *Saccharomyces cerevisiae* supplements vary widely depending on substrate and processing, influencing nutrient content (including B vitamins and selenium) and cell wall composition (including beta-glucans).[24–26] Moreover, yeast beta-glucans (beta-1,3 backbone with beta-1,6 branches) are thought to act largely via immune pathways[27, 28], not to be confused with cereal beta-glucans (mixed beta-1,3/1,4 linkages) which primarily affect gut metabolic processes and glucose homeostasis.[29] Because *Saccharomyces cerevisiae* contains ∼9–18% mixed β-1,3/1,6-glucans,[30] any lactation effect from brewers’ yeast could be from β-glucan itself or from other yeast constituents. However, our findings suggest it may be unlikely that any beneficial effect of brewers’ yeast is related to beta-glucan content directly.

Despite no clear differences between treatment groups, the 95% confidence interval for the treatment effect of brewers’ yeast includes differences that are clinically meaningful. For example, the observed mean difference of 90 mL/day in those taking brewers’ yeast compared with placebo is similar to the 88 mL/day difference shown with the use of the pharmaceutical galactagogue domperidone, which is widely prescribed to mothers in clinical practice for treating or prevention lactation insufficiency.[31] Our findings support the need for a larger adequately powered trial, with potentially longer treatment intervention period, to provide definitive evidence of the efficacy of brewers’ yeast for increasing breast milk production. This is backed up by potentially promising findings of a higher rate of exclusive breast milk feeding on infant discharge in those taking brewers’ yeast compared with placebo (RR 1.32; 95% CI 0.98, 1.78) which warrant further evaluation.

Strengths of this study include employing a rigorous multicentre, double-blind, randomised design and use of an objective robust primary outcome measure. We also examined a comprehensive range of maternal and infant outcomes, extending follow-up until infant discharge or term corrected age, capturing both early and clinically relevant longer-term breastfeeding measures. Limitations include comparatively small sample size per arm, as well as modest levels of adherence to assigned supplements and optimal milk expression frequency, potentially obscuring modest but clinically meaningful differences. The seven-day intervention period may also have been too brief to fully assess efficacy.

## CONCLUSION

We found inconclusive evidence that short-term administration of brewers’ yeast or beta-glucan following preterm birth improves breast milk production, while both interventions appear well tolerated. Given imprecise risk estimates, these findings do not rule out the possibility of a clinically meaningful benefit of brewers’ yeast and suggest further research with a larger sample size may be warranted to clarify the potential clinical impact.

## Data Availability

Data are available upon reasonable request.

## Acknowledgements

The authors wish to acknowledge support provided by the National Health and Medical Research Council Centre of Research Excellence in Optimising human milk nutrition to improve the long-term health of preterm infants (GNT 2024589). The authors also wish to acknowledge Dr Emma Knight for her assistance in providing statistical support with respect to sample size calculation, randomisation, and statistical analysis methods, and Dr Jennie Louise for assistance in supervising the statistical analysis. The authors also wish to acknowledge the Clinical Trials Staff involved with participant recruitment and data collection, including Caitlin Rohal, Laura Cimarosti, Tamara Varcoe, and Grace Holland.

## Contributors

Conceptualised and designed the study: LEG and LHA. Obtained grant funding to conduct the study: LEG, LHA, AK, and ARR. Drafted the manuscript: BS and LEG. Revised the manuscript: LW, ARR, BS, RLK, LY, KD, WVI, AK, KAM, LHA. All authors read and approved the final manuscript. All authors approved the final manuscript as submitted and agreed to be accountable for all aspects of the work. LEG is responsible for the overall content as guarantor.

## Patient consent for publication

Not applicable.

## Ethics approval

This study obtained ethics approval from the Women’s and Children’s Health Network Human Research Ethics Committee, Women’s and Children’s Hospital 72 King William Road North Adelaide SA, Australia (HREC/22/WCHN/00001). Informed consent was obtained from all individual participants included in the study.

## Provenance and peer review

Not commissioned; externally peer reviewed.

## Data availability statement

Data are available upon reasonable request.

## Supplemental material

(added by journal)

