## Supplemental Appendix 1 for "Effect of brewers’ yeast or beta-glucan derived from *Saccharomyces cerevisiae* on breast milk supply following preterm birth: The BLOOM randomised controlled trial"

### Can Brewer's yeast or beta-gLucan increase mOthers Own Milk supply following preterm birth?

STUDY IDENTIFIER: BLOOM

VERSION: 1

DATE: 15/06/2022

PRINCIPAL INVESTIGATOR:
A/Prof Luke Grzeskowiak

SPONSORING INSTITUTION:
South Australian Health and Medical Research Institute
SAHMRI Women and Kids
72 King William Road, North Adelaide SA 5006, AUSTRALIA

Australian and New Zealand Clinical Trials Registry: ACTRN12622000968774

**PROTOCOL AMENDMENTS**

| Version | Date of HREC Approval | Amendments |
|---|---|---|
| V1 | | |

**STATEMENT OF COMPLIANCE**

This document is a protocol for a clinical research study. The study will be conducted in compliance with all stipulations of this protocol, the conditions of ethics committee approval, the NHMRC National Statement on Ethical Conduct in Human Research (2007 – updated May 2015) and the Note for Guidance on Good Clinical Practice (CPMP/ICH-135/95).

I agree that the study will be conducted in accordance with the conditions outlined in the protocol (subject to any amendments). I have read and understood the protocol.

I understand that the information in this protocol is confidential. Publication of information related to this protocol in formats including, but not limited to, conference abstracts, posters or presentations; seminars, journal articles, public reports and internet postings, must be submitted to the Study Steering Committee for consideration. Proposals for said activities must be within a reasonable time frame of any due dates. Approval for all said activities must have the written permission of the Chair of the Steering Committee or their delegate prior to the event.

**Investigator's Name:**

**Investigator's Signature:**

**Date:**

**Study Site:**

**COORDINATING CENTRE:**

Chair Steering Committee: A/Prof Luke GRZESKOWIAK

South Australian Health and Medical Research Institute

**Chair Steering Committee signature:**

**Date:**

**GLOSSARY OF ABBREVIATIONS**

| | |
|---|---|
| AI | Associate Investigator |
| ANZNN | Australian and New Zealand Neonatal Network |
| AE | Adverse Event |
| AR | Adverse Reaction |
| CI | Chief Investigator |
| CRF | Case Report Form |
| EPDS | Edinburgh Postnatal Depression Scale |
| GCP | Good Clinical Practice |
| GMP | Good Manufacturing Practice |
| HREC | Human Research Ethics Committee |
| MPAS | Maternal Postnatal Attachment Scale |
| NHMRC | National Health and Medical Research Council |
| NEC | Necrotizing Enterocolitis |
| NICU | Neonatal Intensive Care Unit |
| PSS:NICU | Parental Stressor Scale: NICU |
| SAE | Serious Adverse Event |
| SAR | Serious Adverse Reaction |
| SAP | Statistical Analysis Plan |
| STAI-6 | Spielberger State-Trait Anxiety Inventory |
| TGA | Therapeutic Goods Association |
| UAR | Unexpected Adverse Reaction |

### 1. SYNOPSIS

| | |
|---|---|
| **Title** | Use of Brewer's yeast or beta-glucan for Optimising mothers Own Milk supply following preterm birth |
| **Acronym** | BLOOM |
| **Objectives** | To determine if routine administration of brewer's yeast or beta-glucan improves breast milk production following preterm birth |
| **Design** | Multicentre, double-blind, randomised controlled trial |
| **Population** | Mothers of preterm infants born at less than 34 weeks' gestation who intend to provide breast milk for their infant and recruited less than 72 hours postpartum. |
| **Outcomes** | **Primary:** Daily breast milk volume on Day 7 of intervention<br>**Secondary:** Adverse events, administration of supplemental feeds (donor milk or infant formula), maternal serum prolactin levels, breast milk composition, breastfeeding rates after treatment completion and at infant discharge. |
| **Study Duration** | The study will be completed within a 2-year period. Participants will receive treatment for 7 days. The primary outcome will be evaluated on day 7. Participants and their infants will be followed until the infant reaches term corrected age or is discharged home from the neonatal unit (whichever occurs first). |
| **Interventions** | **Groups:** Brewer's yeast (1680 mg/day)<br>Beta-glucan (250 mg /day)<br>Placebo<br>**Route:** Enteral<br>**Duration:** 7 days |
| **Sample size** | 99 women (33 in each arm) |

### 1. INVESTIGATORS AND FACILITIES

#### 1.1 *Study Investigators*

**A/Prof Luke E GRZESKOWIAK**
Flinders Medical Research Institute
Flinders University of South Australia
Bedford Park SA 5042
AUSTRALIA
&
SA Pharmacy, Flinders Medical Centre
SA Health
Bedford Park SA 5042


*Authorised to sign study protocol and amendments*

**Prof Lisa AMIR**
Judith Lumley Centre
La Trobe University
Bundoora Campus VIC 3086
AUSTRALIA
&
The Royal Women's Hospital
Parkville VIC 3052
AUSTRALIA


*Breastfeeding medical expert for trial*

**A/Prof Alice RUMBOLD**
SAHMRI Women and Kids
Women's and Children's Hospital
North Adelaide SA 5006
AUSTRALIA

**A/Prof Wendy INGMAN**
Adelaide Medical School
University of Adelaide
Woodville SA 5011
AUSTRALIA

**Dr Amy KEIR**
Neonatal Medicine
Women's and Children's Hospital
North Adelaide SA 5006
AUSTRALIA

**Dr Tina BIANCO-MIOTTO**
School of Agriculture Food & Wine
University of Adelaide
Adelaide SA 5005
AUSTRALIA

#### 1.2 *Principal Site Investigators*

**Dr Amy KEIR**
Neonatal Medicine
Women's and Children's Hospital
North Adelaide SA 5006
AUSTRALIA


**Dr Scott MORRIS**
Dept Perinatal Medicine Flinders Medical Centre Bedford Park SA 5042
AUSTRALIA


#### 1.3 *Statisticians*

**Dr Emma KNIGHT**
SAHMRI Women and Kids
Women's and Children's Hospital
North Adelaide SA 5006
AUSTRALIA


#### 1.4 *Study Locations*

**Women's and Children's Hospital**
North Adelaide SA 5006
AUSTRALIA

**Flinders Medical Centre**
Bedford Park SA 5042
AUSTRALIA

### 2. STUDY MANAGEMENT

The South Australian Health and Medical Research Institute is the nominated sponsor for the trial.

The Principal Investigator at each study centre will be responsible for the conduct of the study at their centre including informed consent, recruitment, data collection and maintenance of study documentation. Handling of investigational products will be the responsibility of a clinical trial staff.

The Coordinating Centre Steering Committee will provide direct day-to-day management for the trial. The core Steering Committee will consist of CIs (Grzeskowiak, Amir, Rumbold) and coordinating centre staff including, but not limited to:

- Senior Trials Manager
- Trial Statistician
- Administrator/Data Manager

The core Steering Committee will meet regularly (at least bi-monthly). Every 3-4 months the Clinical Investigators' Group (CIG) will meet. This will compromise all co-applicants and members of the core Steering Committee.

### 3. SERIOUS ADVERSE EVENT AND TRIAL MONITORING COMMITTEES

#### 3.1 *Serious Adverse Event Committee*

A Serious Adverse Event (SAE) Committee blinded to the treatment allocation will be established and chaired by CI Grzeskowiak and will include medical practitioners with expertise in obstetrics and neonatology. The primary role of the SAE Committee is to review all maternal and infant SAEs to determine whether there is any likelihood that involvement in the trial could have contributed. Cause of death will be determined from autopsy results or other hospital summaries by relevant medical personnel. This committee will meet six-monthly (or as required).

#### 3.2 *Data and Safety Monitoring Committee*

An independent Data and Safety Monitoring Committee (DSMC) will be set up to review the yearly progress of the trial and provide feedback to the Steering Committee. The DSMC will review general study progress (recruitment, compliance, loss to follow-up), breast milk supply and key secondary/safety outcomes, and will have access to unblinded data. The DSMC will also provide advice regarding external issues that may impact on the study (for example changes in clinical practice). This committee will meet yearly or as required and will consist of a:

- Neonatologist/Obstetrician
- Clinical Pharmacologist
- Biostatistician

### 4. FUNDING

This study is supported by funding received from Leiber GmbH, Germany as well as well as the Channel 7 Children's Research Foundation, South Australia, Australia.

### 5. INTRODUCTION AND BACKGROUND

#### *Mothers' own breast milk is best*

Breast milk is considered the optimal nutrition for preterm infants.[1, 2] A compelling body of evidence shows breast milk reduces neonatal morbidity and mortality.[3] Use of mothers' own breast milk during hospitalisation reduces the incidence and severity of preventable morbidities, including necrotizing enterocolitis (NEC), late onset sepsis, chronic lung disease, retinopathy of prematurity, rehospitalisation after discharge, and neurodevelopmental problems in infancy and childhood.[3] For example, a recent Cochrane review showed preterm infants receiving infant formula are three times more likely to develop NEC, which has a mortality rate of 20-40%.[4] Further, the ability for a mother to provide her own breast milk represents an important tangible contribution in a situation where mothers are often unable to provide much infant care in these first few weeks of life. The resultant psychological benefits include greater feelings of attachment, empowerment, and confidence.[5]

#### *Physiology of lactation*

Lactation is regulated by several reproductive (oestrogen, progesterone, placental lactogen, prolactin, and oxytocin) and metabolic hormones (glucocorticoids, insulin, growth, and thyroid). Reproductive hormones act directly on the mammary gland, whereas metabolic hormones have indirect actions through altering endocrine responses and nutrient transfer to the mammary gland.

Development of functional lactation is a multi-stage event. Secretory differentiation (lactogenesis I) occurs in mid- to late pregnancy when differentiation of mammary epithelial cells into lactocytes occurs, conferring the ability to synthesize and secrete key components of human milk. Secretory activation (lactogenesis II) is triggered by the sudden drop in progesterone following delivery of the placenta, accompanied by high levels of circulating prolactin. Milk secretion is copious and occurs between 24–102 hours (average 60 hours) after the birth. This is often described as the milk 'coming in.' Oxytocin is released in response to infant suckling, stimulating contraction of myoepithelial cells around the alveoli and facilitating milk release.[6] Apart from a sudden increase in breast milk volume, the process of secretory activation can be identified by a rapid increase in lactose, citrate, and sodium concentrations in the breast milk. Once lactation is established, an autocrine control (local feedback) mechanism regulates ongoing milk production, meaning supply is largely based on effective removal of milk from the breast.[6]

#### *Preterm birth and insufficient breast milk supply*

Mothers of preterm infants face many challenges in establishing and maintaining an adequate supply of breast milk during their infant's prolonged hospitalisation. This is driven by multiple factors including; physiological immaturity of the breast associated with preterm birth, inability for the preterm infant to breastfeed directly from the breast, and the stress of having an infant admitted to the Neonatal Intensive Care Unit (NICU). Each of these factors has the ability to interfere with establishment of normal milk supply; one previous study demonstrated 82% of women birthing preterm experienced delayed secretory activation.[7] While longer-term breastfeeding outcomes were not collected for this cohort, other studies in mothers of term infants have demonstrated that delayed secretory activation is associated with increased risk of early cessation of breastfeeding.[8]

Some mothers may respond well to non-pharmacological lactation support strategies (i.e. correct use of breast milk pump, increasing expressing frequency) for increasing breast milk supply, but for a large number of mothers, their breast milk supply continues to be insufficient to meet their infant's needs. Insufficient volume of mothers' own breast milk has been partly addressed through the introduction of human milk banks, however there are only five banks across Australia. Further, mothers' own milk has been demonstrated to be superior to donor human milk with respect to composition and bioactivity, highlighting that focusing on supporting mothers to provide their own breast milk to their infants is key to optimising neonatal outcomes.[3]

Given the challenges mothers of preterm infants face with respect to initiation and sustaining lactation, attention has shifted towards the potential role of early non-pharmacological and pharmacological interventions in improving breast milk supply soon after birth. A recent survey of 1876 Australian women found that 60% reported taking galactagogues, known as substances thought to aid in initiating and maintaining adequate milk production, during breastfeeding.[9] This is despite a lack of clear evidence to guide their use.[10-11] The most commonly reported galactagogues in the survey included lactation cookies and brewer's yeast.[9] While lactation cookies may vary in composition, they typically consist of ingredients such as oats, brewer's yeast and flaxseed. The active ingredient of oats is thought to be beta-glucan, which is a glucose polymer present in the cell walls of cereals as well as yeast (i.e. brewers' yeast) and fungi.[12]

While natural galactagogues, such as brewer's yeast, are widely perceived by women to be safer than pharmaceutical galactagogues and are taken by many women,[9] evidence to support their efficacy is largely absent. With respect to brewers' yeast, the likely mechanism of action remains unknown. Some have postulated that it could relate to the high concentration of B vitamins, or the presence of beta-glucan isolated from the cell wall.[13] Studies based on animal models provide evidence that intravenous injection of beta-glucan leads to a significant increase in serum prolactin,[14] providing a potential pathway to changes in breast milk production. The relevance of such findings to oral consumption of beta-glucan in humans, however, remains unclear given the limited bioavailability. In contrast, a number of studies have demonstrated that oral consumption of beta-glucans may have immunomodulatory effects.[12] As immune dysregulation is a common feature of preterm birth[15] and has been associated with impaired lactation in animal models[16], this represents an alternative potential mechanism of action in which brewers' yeast/ beta-glucans could influence breast milk production. Data in lactating women, however, remains scant. A recent clinical trial by Wesolowska et al evaluated the efficacy of a barley malt-based galactagogue (containing a proprietary blend of barley malt and beta-glucan) compared with placebo in mothers of preterm infants.[17] Compared with placebo, those who took the barley malt preparation reported expressing a greater total volume of breastmilk over the 14 day intervention period (6036 ± 498 vs 4209 ± 335; p=0.003). Differences in daily expressed breast milk volume between groups were evident by day 7 of the intervention. No women reported experiencing any side effects during the study. However, the study can only be considered to provide low quality evidence due to a substantial loss to follow-up of 32% across both treatment groups. Therefore, it remains unknown whether natural galactagogues such as brewers' yeast or beta glucan are effective in improving breast milk production following preterm birth.

##### *SUMMARY*

Mothers of preterm infants often struggle in producing enough breast milk to meet the daily feed requirements of their infants, especially in the longer-term. This randomized controlled trial will evaluate the efficacy and safety of two commonly used galactagogues, Brewer's yeast and beta-glucan, compared with placebo in improving maternal breast milk supply following preterm birth.

### 6. STUDY OBJECTIVES

#### 6.1 *Primary objective*

To determine if routine administration of brewer's yeast or beta-glucan improves breast milk production following preterm birth.

### 7. STUDY DESIGN

#### 7.1 *Type of study*

Multicentre, double-blind, randomised controlled trial. Participants, care providers, outcome assessors, trial investigators, and data analysts will be blinded to randomisation group.

#### 7.2 *Number of participants*

The planned sample size is a total of 99 women who gave birth to preterm infants at less than 34 weeks' completed gestation (i.e. 33 per randomised group)

#### 7.3 *Expected duration*

The study will be completed within a 2-year period. Participants will receive treatment for 7 days when the primary outcome will be evaluated. Participants and their infants will be followed until the infant reaches term corrected age or is discharged home from the neonatal unit (whichever occurs first).

##### 7.4 *Primary outcome measures*

Daily breast milk volume on Day 7 following randomisation.

##### 7.5 *Secondary outcome measures*

Adverse events, administration of supplemental feeds (donor milk or infant formula), maternal serum prolactin levels, breast milk composition, breastfeeding rates after treatment completion at infant discharge.

#### 8. STUDY TREATMENTS

##### 8.1 *Treatment arms*

Participants will be randomised to one of three treatment arms, consisting of either brewer's yeast (*Saccharomyces cerevisiae*), beta-glucan (purified from *Saccharomyces cerevisiae*) or placebo (which will be identical to investigational product in appearance).

##### 8.2 *Dosage and route of administration*

All study participants will take three capsules twice daily (six capsules a day) according to the treatment schedule outlined below.

Brewer's Yeast: 1680 mg / day

Beta-glucan: 250 mg / day

Placebo: Microcrystalline cellulose

| **Table 1. Dosing regimen** | | |
|---|---|---|
| **Treatment Arm** | **Morning** | **Night** |
| Brewers' Yeast | 3 x 280 mg capsules | 3 x 280 mg capsules |
| Beta-glucan | 2 x placebo capsules<br>1 x 250 mg capsule | 3 x placebo capsules |
| Placebo | 3 capsules | 3 capsules |

To facilitate treatment blinding and aid medication adherence, the medications will be provided in bottles with instructions according to dosing regimen above . All study medications will be identical in appearance.

##### 8.3 *Description*

Study medications will be provided in the form of capsules, identical in appearance.

#### 8.4 *Manufacturer*

Study medications will be manufactured and supplied by Leiber GmbH (Germany).

#### 8.5 *Packaging and labelling*

Participants will be provided sufficient capsules for 7 days of treatment. Each investigational product will be identical in appearance, weight and packaging. The study medications will be packaged and labelled in accordance with GMP including product ID, batch number, expiry date and include the statement "for clinical trial use only".

#### 8.6 *Medication storage*

Study medications will be stored in a designated secured area at each site and clearly labelled for research purposes only. An inventory will be kept of study medication supplies at all sites.

#### 8.7 *Dispensing and product accountability*

The study coordinator will maintain accurate records of the receipt of all study medication, and when and how much study medication is dispensed and used by each participant in the study. Reasons for departure from the expected dispensing regimen will be recorded.

At the end of the study, there will be final reconciliation of study medications received, dispensed, consumed and returned.  Any discrepancies will be investigated, resolved and documented by the study team. Unused study medications will be destroyed in compliance with applicable regulations.

#### 8.8 *Medication adherence*

Medication adherence will be determined informally at the end of the study by the Principal Investigator (PI) of each centre (or nominee), incorporating pill counts. In addition, women will also be provided with a medication diary to keep track of all doses taken (or missed).

#### 8.9 *Concomitant medications/treatment*

Given the absence of any known drug-drug interactions, there are no restrictions on concomitant medications.

### 9. ENROLMENT AND RANDOMISATION

Eligible participants will be provided information on the study by their nurse or midwife who will ask them to complete a Consent to Contact form. Upon providing consent, SAHMRI study staff will contact potential participants to explain the study.  The information sheet will describe the purpose of the study, the procedures to be followed, and the risks and benefits of participation. Study staff will conduct the informed consent discussion and will confirm that information provided is understood and answer any questions about the study. Consent will be voluntary and free from coercion.  A copy of the consent form will be given to participants and documented in their infant's medical record and study CRF. When all the inclusion and exclusion criteria have been addressed and the eligibility of the participant confirmed, the participant will be randomly assigned to one of three treatment arms.

A record of all mothers screened but not enrolled will be maintained.

#### 9.1 *Inclusion criteria*

Each participant must meet all the following criteria to be enrolled in this study:

- Infant born <34 weeks' gestation (i.e. up to 33+6)
- Intention to provide breast milk
- Between 0 to 72 hours of birth
- Age ≥ 18 years
- Willing and able to comply with all study requirements, including treatment, timing and/or nature of required assessments
- Adequate English language skills
- Signed, written informed consent

#### 9.2 *Exclusion criteria*

- Contraindication to breastfeeding (i.e. HIV)
- Higher order pregnancies (triplet or more)

#### 9.3 *Randomisation procedures*

Women will be allocated to receive either brewer's yeast, beta-glucan, or placebo.

Participants will be randomised using the REDCap Randomisation Module. Participants will be randomised to one of the three treatment arms in a 1:1:1 ratio. The randomisation schedule will be prepared using ralloc.ado in Stata by an independent statistician who is not involved with trial participants or data. The randomisation schedule will be stratified according to study centre.

The schedule will be kept by the independent statistician and the treatment allocation of each randomisation code can be provided to the investigator in case of emergency.

Women must be randomised before starting study treatment. Treatment should be started as soon as possible and preferably within 24 hours of randomisation. Randomisation should occur only after all screening assessments have been performed, participant's eligibility verified and signed consent obtained. Once the randomisation process has been completed as per the instructions in the Study Manual, the woman will be assigned a treatment arm and a study ID.

#### 9.4 *Blinding*

Participants and their care providers, outcome assessors, trial investigators and data analysts will be blinded to randomisation group.

#### 9.5 *Breaking of the study blind*

##### 9.5.1 On study

The randomisation code for an individual participant may only be unblinded in emergency situations, where the Investigator decides a participant cannot be adequately treated without knowing the identity of their treatment allocation. The Principal Investigator must be contacted. All attempts to avoid breaking the code (i.e. withdrawal of treatment) should be made. To break the randomisation code the Investigator must contact the randomisation facility/personnel. The time, date, participant study ID and reason for unblinding must be documented. Events leading to the emergency breaking will be recorded in the serious adverse event (SAE) report form.

##### 9.5.2 Following completion of the study

Trial allocation codes will only be unblinded once all data collected has been entered into the study database for every participant, the database has been finalised and analysis of primary outcome using blinded treatment codes has been completed, except in the case of an emergency, as detailed above.

#### 9.6 *Treatment discontinuation*

Study treatment will be permanently discontinued for any of the following reasons:

- Unacceptable toxicity as determined by the participant or site investigator
- The investigator determined that continuation of treatment is not in the participant's best interest
- Failure to comply with the protocol, the participant declines further study treatment, or withdraws their consent to participate in the study.

#### 9.7 *Participant withdrawal*

Women are free to withdraw themselves and/or their infant from the study at any time. The reasons for withdrawal will be recorded in the CRF and included in the final report. Participants who discontinue treatment or are withdrawn from the study will not be replaced. Whenever possible, permission will be sought from participants who withdraw from the study to obtain as much data for the follow-up period as they will permit.

### 10. STUDY ASSESSMENTS AND PROCEDURES

#### 10.1 *Study procedures*

Participants in the study will be asked to partake in a total of four study related appointments. Appointments will be conducted within the hospital (either within the neonatal unit, postnatal ward, or designated clinical research area), or by telephone (where appropriate).

##### 10.1.1 Demographic/Lifestyle Questionnaire

Background information will be collected in order to describe the characteristics of the study cohort sample. Baseline maternal demographic and lifestyle characteristics include age, height, pre-pregnancy weight, pregnancy weight gain, smoking status, alcohol intake, pre-existing medical conditions (e.g. polycystic ovarian syndrome, diabetes, thyroid disorders), pregnancy complications (e.g. gestational diabetes) and medication use.

##### 10.1.2 Breast Milk Diary

Participants will be asked to maintain a daily breast milk diary throughout each day of the study. This diary will record the number of times each breast was expressed, the method used to express, the volume of expressed milk and if applicable information pertaining to direct breastfeeding. Women will be asked to continue using the diary for 21 days. If the infant directly breastfeeds on day 7 of the study, then test-weighing (i.e. infant weight before and following breastfeed) will be used to estimate breast milk intake.

##### 10.1.3 Breast Health and Milk Expression Questionnaire

Participants will be asked questions relating to methods of breast expression from birth to study enrolment as well as perceived onset of secretory activation. During the study, women will be asked to report data on general breast health (e.g. covers symptoms associated with mastitis), including the 'Breast milk expression experience measure' questionnaire.

##### 10.1.4 Postnatal Health Questionnaire

Participants will be asked to detail postnatal health issues during the study (e.g. infections, cold/flu), as well as any medications (prescription or non-prescription), herbal supplements or multivitamins taken.

##### 10.1.5 Mental Health and Wellbeing Questionnaires

Maternal depressive and anxiety symptoms will be collected using standardised validated questionnaires (Edinburgh Postnatal Depression Scale [EPDS], Spielberger State-Trait Anxiety Inventory [STAI-6]). Stress related to infant hospitalisation will be assessed using the Parental Stressor Scale: NICU (PSS:NICU). Maternal-infant attachment will be assessed using the Maternal Postnatal Attachment Scale (MPAS).

Responses to the EPDS will be managed according to our Standard Operating Procedure. In brief, women identified as being at high risk of depression based on responses to the EPDS (i.e. a score of ≥ 13 or positive response to Q10 regarding self-harm) will be notified by study staff of the result, offered written information regarding access to mental health support services and asked to provide consent to share the findings with a trusted healthcare professional in order to obtain further help.

##### 10.1.6 Infant Feeding Practices Questionnaire

Women will complete an infant-feeding questionnaire to determine whether they are breastfeeding exclusively, using a combination of breast- and formula feeding, or formula feeding only.

##### 10.1.7 Medical Record Audit

Maternal and infant case notes will be reviewed to collect data on pregnancy history (e.g. gestational diabetes) and labour/delivery outcomes (e.g. method of delivery, receipt of antenatal steroids prior to delivery, receipt of magnesium sulphate prior to delivery). Infant details collected from medical records include birth weight, gestational age, birth length, head circumference, 1- and 5-minute APGAR scores.

##### 10.1.8 Adverse Events

Information pertaining to potential adverse events or serious adverse events will be collected at each study visit until seven days post-trial completion. Women will also be encouraged to write down any adverse events experienced during the trial in their breast milk production diary.

##### 10.1.9 Medication Compliance Check

Women will be supplied a dosing schedule to record each dose that is taken throughout the study. Further, at the end of each week women will be asked whether they remembered to take all their study medications to evaluate compliance. On day 7 of the study, women will be asked to bring in the study medication packaging, with any remaining capsules to be counted.

##### 10.1.10 Maternal Anthropometrics

Maternal weight, waist circumference, hip circumference, and mid upper-arm circumference will be assessed to evaluate changes in body composition related to the study intervention.

##### 10.1.11 Random Blood Sample

A non-fasted blood sample will be collected to evaluate changes in prolactin, as well as cardiometabolic and inflammatory biomarkers. Blood samples will also be used to obtain maternal DNA. This will be used to perform genetic and epigenetic tests to identify differences in gene variants, DNA methylation and telomere length associated with breast milk supply and galactagogue treatment.

##### 10.1.12 Breast Milk Sample

Breast milk samples (8 mL) will be collected to assess changes in macronutrient composition. A sample will be taken from breast milk expressed between an agreed specified time (9am-11am) in accordance with our Standard Operating Procedure.

###### 10.1.13 Maternal Urine Sample

A urine sample will be collected to assess urinary metabolites and environmental toxins such as phthalates. The sample will be collected, processed and stored in accordance with our Standard Operating Procedures.

###### 10.1.14 Maternal Stool Sample

A stool sample will be collected to assess the maternal microbiome in relation to the breast milk and infant microbiomes. The sample will be collected, processed and stored in accordance with our Standard Operating Procedures.

###### 10.1.15 Maternal Buccal Swab

A buccal swab will be collected to investigate differences in DNA methylation according to breast milk supply and treatment response to brewer's yeast or beta-glucan. The sample will be collected, processed and stored in accordance with our Standard Operating Procedures.

###### 10.1.16 Infant Growth and Anthropometric Measures

Infant head circumference, length and weight measured weekly by medical staff during the infants' hospitalisation will be collected from the infant medical record.

###### 10.1.17 Infant Inpatient Feeding Record (from medical record audit)

The number of days taken to reach full enteral feeds (enteral intake ≥120 mL/kg/day for 3 consecutive days), day of age feeds commenced, days on parenteral nutrition, days on intravenous lipids and type of lipids, type of milk at feed commencement, and at discharge home will be collected from the infant medical record.

###### 10.1.18 Infant Morbidity/Mortality* (from medical record audit)

The number of days in hospital (to first discharge home), postnatal steroid use, grade of intraventricular haemorrhage (IVH), confirmed sepsis, confirmed necrotizing enterocolitis (NEC), grade of retinopathy of prematurity (ROP), surgical procedures and death during first hospitalisation. This data will be collected from the infant medical record.

*All infant clinical data will be collected in accordance with the definitions of the Australian and New Zealand Neonatal Network (ANZNN).[22]

#### 10.2 *Study Assessments*

##### 10.2.1 Study Visit 1 – Randomisation Appointment (Day 0)

<u>Maternal assessments</u>

- Assess study eligibility
- Randomise to treatment group
- Commence Daily Milk Production Diary (10.1.2)
- Medical Record Audit – Pregnancy History (10.1.7)

- Demographic/Lifestyle Questionnaire (10.1.1)
- Mental Health and Wellbeing Questionnaires (10.1.5)
    - EPDS; STAI-6

##### 10.2.2 Study Visit 2 (Day 7)

*Protocol window – day 7-9*

Maternal assessments

- Breast Milk Production Diary Check (10.1.2)
- Mental Health and Wellbeing Questionnaires (10.1.5)
    - EPDS; STAI-6; PSS-NICU
- Breast Health Questionnaires (10.1.3)
- Adverse Event Assessment (10.1.8)
- Medication Compliance Check (10.1.9)
- Breast Milk Sample (10.1.12)
- Maternal Urine Sample (10.1.13)
- Maternal Stool Sample (10.1.14)
- Maternal Buccal Swab (10.1.15)
- Maternal Blood Sample (10.1.11)

##### 10.2.3 Study Visit 3 (Day 21 postpartum)

*Protocol window – day 21-23*

Maternal assessments

- Breast Milk Production Diary Check (10.1.2)
- Breast Health Questionnaires (10.1.3)
- Adverse Event Assessment (10.1.8)
- Postnatal Health Questionnaire (10.1.4)

##### 10.2.4 Study Visit 4 (Infant at discharge or term corrected age)

*Protocol window – within 3 days of event*

Maternal assessments

- Infant-Feeding Practices Questionnaire (10.1.6)
- Mental Health and Wellbeing Questionnaires (10.1.5)
    - EPDS; STAI-6; PSS-NICU; MPAS

##### 10.2.5 Medical Record Audit (post-discharge)

- Infant Feeding Data (10.1.17)
- Infant Morbidity/Mortality (10.1.18)
- Infant Growth and Anthropometric Measures (10.1.16)

#### 10.3 *Schedule of assessments – maternal*

| MATERNAL | INTERVENTION PHASE | | | FOLLOW-UP PHASE | |
|---|---|---|---|---|---|
| | Screening | Baseline | Day 7 | Day 21 Postpartum | Infant Discharge to Home or Term Corrected |
| **General** | | | | | |
| Eligibility assessment | X | | | | |
| Informed consent | X | | | | |
| Randomisation | | X | | | |
| **Questionnaires** | | | | | |
| Demographic/Lifestyle questionnaire | | X | X | | X |
| Breast milk diary (daily breast milk volume) | | X | X | X | |
| Infant-feeding practices questionnaire | | | | | X |
| Mental health and wellbeing questionnaires<br>- EPDS, STAI-6 | | X | X | | X |
| - PSS:NICU | | | X | | X |
| - MPAS | | | | | X |
| Breast health questionnaire | | | X | X | X |
| **Physical Assessments** | | | | | |
| Anthropometric measurements | | | X | | |
| **Case Note Audit** | | | | | |
| Pregnancy/Birth history | | X | | | |
| **Biospecimens** | | | | | |
| Collection of untimed blood for research# | | | X | | |
| Collection of breast milk for research# | | | X | | |
| Collection of urine for research# | | | X | | |
| Collection of stool for research# | | | X | | |
| Collection of buccal swab for research# | | | X | | |

*# Biological samples to be collected at selected sites only. Refer to Standard Operating Procedure for collection procedures.*

#### 10.4 *Schedule of assessments – infant*

| | INTERVENTION PHASE | | | FOLLOW-UP PHASE | |
|---|---|---|---|---|---|
| | Screening | Randomisation / Baseline | Day 7 | Day 21 postpartum | Infant Discharge to Home or Term Corrected |
| **Case Note Audit** | | | | | |
| Infant feeding (daily feeding data) | | X------------------- | ------------------- | ------------------- | -------------------\| |
| Infant morbidity during admission (ANZNN Registry) | | | | | X |
| Anthropometric measurements (as part of routine care) | | X | X | X | X |

### 11. ADVERSE EVENT REPORTING

Any unfavourable and unintended sign, symptom or illness that develops or worsens during the period of the study will be classified as an adverse event (AE), whether it is considered to be related to the study treatment. Adverse events will include unwanted side effects, sensitivity reactions, abnormal laboratory results, injury or inter-current illnesses, and may be expected or unexpected. These will be recorded electronically on the CRF.

Safety evaluations will be conducted each week during the study. Study PI or site-PIs can be directly contacted by the participants if there are any concerns regarding their treatment. The period for adverse event reporting will be from the time of first dose until seven days post final study medication administration. The participants will be followed up face-to-face or by telephone interview at twenty-one days post-partum.

#### 11.1 *Safety reporting for RCT*

Definitions of harm of the EU Directive 2001/20/EC Article 2 based on the principles of ICH GCP apply to this trial.

**Table 2: Adverse Event Definitions**

| | |
|---|---|
| Adverse Event (AE) | Any untoward medical occurrence in a patient or clinical trial participant administered a medicinal product and which does not necessarily have a causal relationship with this product. |
| Adverse Reaction (AR) | Any untoward and unintended response to an investigational medicinal product related to any dose administered |
| Unexpected Adverse Reaction (UAR) | An adverse reaction, the nature or severity of which is not consistent with the applicable product information (eg Investigator's Brochure for an unauthorised product or summary of product characteristics (SPC) for an authorised product. |
| Serious Adverse Event (SAE) or Serious Adverse Reaction (SAR) | Any AE or AR that at any dose:<br>• results in death<br>• is life threatening*<br>• requires hospitalisation or prolongs existing hospitalisation**<br>• results in persistent or significant disability or incapacity<br>• or is another important medical condition*** |
| * The term life threatening here refers to an event in which the patient is at risk of death at the time of the event; it does not refer to an event that might hypothetically cause death if it was more severe (eg a silent myocardial infarction) | |

| |
|---|
| ** Hospitalisation is defined as an in-patient admission, regardless of length of stay, even if the hospitalisation is a precautionary measure for continued observation. Hospitalisation for pre-existing conditions (including elective procedures that have not worsened) do not constitute an SAE<br><br>*** Medical judgement should be exercised in deciding whether an AE or AR is serious in other situations. Important AEs or ARs that may not be immediately life threatening or result in death or hospitalisation, but may seriously jeopardise the participant by requiring intervention to prevent one of the other outcomes listed in the table (eg a secondary malignancy, an allergic bronchospasm requiring intensive emergency treatment, seizures or blood dyscrasias that do not require hospitalisation, or development of drug dependency). |

**Adverse events include:**

- an exacerbation of a pre-existing illness
- an increase in the frequency or intensity of a pre-existing episodic event or condition
- a condition (regardless of whether PRESENT prior to the start of the trial) that is DETECTED after trial drug administration. (This does not include pre-existing conditions recorded as such at baseline – as they are not detected after trial drug administration.)
- continuous persistent disease or a symptom present at baseline that worsens following administration of the trial treatment

**Adverse events do NOT include:**

- Medical or surgical procedures: the condition that leads to the procedure is the adverse event
- Pre-existing disease or a condition present before treatment that does not worsen
- Hospitalisation where no untoward or unintended response has occurred (eg elective cosmetic surgery)
- Overdose of medication without signs or symptoms

#### 11.2 *Seriousness assessment*

When an AE or AR occurs, the investigator responsible for the care of the participant must first assess whether the event is serious. For infant AEs, any deaths will be classified as serious. For maternal AEs, seriousness will be assessed using the definition given in Table 2. If the event is classified as 'serious' then an SAE form must be completed and the Chair of the Steering Committee (or delegated body) notified within one working day.

#### 11.3 *Serious Adverse Events (SAE)*

Determination of the relevant category for reporting a medical event/reaction in the trial will be conducted according to the safety reporting assessment flow chart depicted in

Figure B, 'Safety monitoring and reporting in clinical trials involving therapeutic goods, National Health and Medical Research Council, 2016'. All maternal and infant SAEs are to be reported to the Chair of the Steering Committee, Dr Luke Grzeskowiak within 24 hours of the site becoming aware. Any event that in the opinion of a Principal Investigator may be of immediate or potential concern for a participant's health or well-being will also be reported immediately to the Chair of the Steering Committee. Emergency contacts are listed in section 13.1.

The Serious Adverse Event Committee will review all SAEs. If a mother or infant dies, any post-mortem findings, including histopathology, must be provided to the Coordinating Centre to allow a full independent review.

##### 11.3.1 Severity or grading of adverse events

The severity of all AEs and/or ARs (serious and non-serious) in this trial should be graded using the toxicity grading according to the Common Toxicity Criteria (version 4, 28 May 2009):

1 – Mild
2 – Moderate
3 – Severe
4 – Life threatening
5 – Death

##### 11.3.2 Causality

The SAE Committee must assess the causality of all SAEs in relation to the trial therapy using the definitions in Table 3.

**Table 3: Causality definitions**

| Relationship | Description | Event Type |
|---|---|---|
| Unrelated | There is no evidence of any causal relationship | Unrelated SAE |
| Unlikely to be related | There is little evidence to suggest that there is a causal relationship (eg the event did not occur within a reasonable time after administration of the trial medication). There is another reasonable explanation for the event (eg the participant's clinical condition or other concomitant treatment) | Unrelated SAE |
| Possibly related | There is some evidence to suggest a causal relationship (eg because the event occurs within a reasonable time after administration of the trial medication). However, the | SAR |

| | | |
|---|---|---|
| | influence of other factors may have contributed to the event (eg the participant's clinical condition or other concomitant treatment) | |
| Probably related | There is evidence to suggest a causal relationship and the influence of other factors is unlikely | SAR |
| Definitely related | There is clear evidence to suggest a causal relationship and other possible contributing factors can be ruled out. | SAR |

##### 11.3.3 Expectedness

As it is not expected that the IMP or study protocol will cause any SAEs to mother or infant, all SARs will be classified as unexpected.

#### 11.4 *Emergency contact details*

Dr Luke GRZESKOWIAK – Principal Investigator and Chair of Steering Committee
Flinders Health and Medical Research Institute
Flinders University
Bedford Park SA 5042


Dr Lisa AMIR (Medical Doctor)
Judith Lumley Centre
Level 3, George Singer Building
La Trobe University
Bundoora VIC 3086
AUSTRALIA


#### 11.5 *Australian Therapeutic Goods Association (TGA) and HREC notification*

The Australian TGA requires notification of serious adverse events which are unexpected and deemed to be related to the study treatments. Fatal or life-threatening unexpected adverse events will be notified to the TGA as soon as possible but no later than 7 calendar days after first knowledge by the Coordinating Centre that a case qualifies, followed by an as complete report as possible within 8 additional calendar days. Serious, unexpected adverse events that are not fatal or life-threatening, but deemed related to study treatment shall be filed as soon as possible but no later than 15 calendar days. Serious adverse events that are unrelated to the study treatments shall be included in the end of study report.

The Investigator, or nominee, will also be responsible for reporting any serious adverse events to their Human Research Ethics Committee (HREC) as soon as possible and in any event within 72 hours. In agreeing to the provisions of the protocol, these responsibilities are accepted by the Investigator, or nominee.

If a participant or their infant dies, any post-mortem findings including histopathology, must be provided to the Coordinating Centre.

### 12. STATISTICAL METHODS

#### 12.1 *Sample size estimation*

A sample of 99 women (33 per arm) yields 90% power, 0.025 alpha to show a difference in the mean daily breast milk volume of 150 mL/day (200 mL/day standard deviation) between each of the intervention arms and control, allowing for 10% loss to follow-up. This includes adjustment for a 0.6 correlation between breast milk volume at study entry and breast milk volume on day 7.

#### 12.2 *Statistical Analysis Plan*

A stand-alone Statistical Analysis Plan (SAP) detailing the prespecified analyses to be performed will be produced during the recruitment stage of the study. The SAP will be approved by the Steering Committee prior to any analyses being conducted. Results will be reported according to the CONSORT statement. It is envisaged that the analysis will be undertaken using R and STATA software.

#### 12.3 *Statistical Methods- Outcomes*

The primary analysis will be performed according to the treatment group to which participants were randomised (intention-to-treat principle). A secondary per-protocol analysis will also be performed for each of the primary and secondary outcomes.

The primary outcome of daily breast milk volume on day 7 will be compared between treatment groups using a linear regression. The results will be expressed as a difference in means with a 95% confidence interval and two-sided p-value. Adjustment will be made for baseline breast milk volume and the randomization strata (study centre). A p-value of less than 0.05 will be considered to indicate statistical significance. Analysis of secondary outcomes will use log-binomial regression models for binary outcomes and linear regression models for continuous outcomes with adjustment for stratification variables and other pre-specified prognostic baseline variables. Results will be presented as relative risks and differences in means respectively, along with 95% confidence intervals. Missing data will be addressed using multiple imputation. Sensitivity analyses will also be performed using the original unimputed data.

Planned sub-group analyses of the primary and secondary breastfeeding outcomes include:

(i) Plurality (Singleton vs. twins)
(ii) Parity (Primiparous vs. multiparous)
(iii) Infant gestation at birth ($23^{+0}$–$29^{+6}$ vs. $30^{+0}$–$31^{+6}$ vs $32^{+0}$-$33^{+6}$ weeks' gestation)
(iv) Maternal body mass index (Underweight/Normal weight vs. Overweight/Obese)

Effect modification by each of these factors will be assessed separately by including interaction effects with treatment group in the statistical models.

### 13. DATA MANAGEMENT

#### 13.1 *Data collection*

Data entry and study management will be handled using REDCap. Electronic CRFs will be used, with data collected and stored directly in REDCap. Paper-based CRFs will be available for use where needed.

In order to ensure the accuracy of data collected, representatives from the Coordinating Centre and regulatory authorities will have access to source documents (i.e. mother's or infant's medical records). Confidentiality will be maintained at all times.

#### 13.2 *Data storage*

Paper based CRFs will be stored in a locked office at the study site. Only research staff directly involved in the study will have access to the information.

Electronic forms of data will be collected and stored using REDCap. No confidential data are stored on data entry machines. Each study PI and research staff will be provided with their unique security login. Access to electronic data is granted only to research staff according to specific need. Access is only granted to specific sections of the database(s) and at levels relevant to that person. Only specified staff, using tools within REDCap that track all events, can modify the database.

Transaction logs of the databases to a hard drive on another secure server will be made in accordance with the needs of the project. Web servers and database servers are physically separate.

#### 13.3 *Study record retention*

Original/copies of study documents will be retained at the study site or in archives. Documents will be retained for at least 30 years after study completion in line with the data retention schedules for research involving minors. At the completion of this time documentation will be destroyed using confidential document disposal, by shredding with a commercial grade document shredder. The study electronic data will be stored indefinitely on SAHMRI's secure servers with access only granted to authorised study personnel.

#### 13.4 *Genomic Data Sharing Plan*

It is an international best practice in large-scale genomics research to make deidentified sequencing data publicly available "to facilitate the translation of research results into knowledge, products and procedures that improve human health" (NIH). The Gene Expression Omnibus (GEO) is a public repository hosted by the National Center for Biotechnology Information (NCBI) at the National Institutes of Health (NIH) in the United States. It archives and distributes comprehensive sets of high-throughput functional genomic data submitted by the scientific community. Many scientific journals require proof of GEO accession numbers for study datasets before acceptance of a paper for publication. Some government funding institutions such as the NIH also require investigators to submit a prespecified genomic data sharing plan as a condition of public research funding.

In order to adhere to international best research practice and to ensure eligibility of our study for publication in scientific journals, we plan to archive the fully deidentified sequencing datafiles in GEO (or equivalent database). The sequencing data files prepared for GEO will be free of all identifiers that would permit linkages to individual research participants and variables that could lead to deductive disclosure of the identity of individual subjects, in accordance with the NIH Statement on Sharing Research Data (2003) [See NIH website for further details on data sharing here https://grants.nih.gov/grants/policy/data_sharing/].

### 14. ADMINISTRATIVE ASPECTS

#### 14.1 *Regulatory compliance*

This study will be conducted according to the Note for Guidance on Good Clinical Practice (CPMP/ICH/135/95) annotated with TGA comments (Therapeutic Goods Administration DSEB July 2000) and in compliance with applicable laws and regulations. The study will be performed in accordance with the NHMRC Statement on Ethical Conduct in Research Involving Humans (© Commonwealth of Australia 2007), and the NHMRC Australian Code for the Responsible Conduct of Research (©Australian Government 2007), and the principles laid down by the World Medical Assembly in the Declaration of Helsinki 2008. To this end, no participant will be recruited to the study until all the necessary approvals have been obtained and the participant has provided written informed consent. Further, the investigator shall comply with the protocol, except when a protocol deviation is required to eliminate immediate hazard to a subject. In this circumstance the Principal Investigator and HREC must be advised immediately.

#### 14.2 *Confidentiality*

Participant confidentiality is strictly held in trust by the participating investigators and research staff and their agents. This confidentiality is extended to cover testing of biological samples in addition to the clinical information relating to participants. The study protocol, documentation, data and all other information generated will be held in strict confidence. No information concerning the study or the data will be released to any unauthorised third party, without prior written approval of the coordinating centre. Coordinating Centre and regulatory authorities may inspect all documents and records required to be maintained by the Investigator, including but not limited to, medical records and pharmacy records for participants and their infants in this study subject to individuals having obtained approval/clearance through State/National Governments and HREC as required by local laws. The clinical study site will permit access to such records. Clinical information will not be released without written permission of the subject, except as necessary for monitoring by HREC or regulatory agencies.

#### 14.3 *Independent HREC approval*

This protocol and the informed consent document and any subsequent modifications will be reviewed and approved by the HREC of each study site. A letter of protocol approval by HREC will be obtained prior to the commencement of the study, as well as approval for other study documents subject to HREC review.

#### 14.4 *Modifications of the protocol*

This study will be conducted in compliance with the current version of the protocol. Any change to the protocol document or Informed Consent Form that affects the scientific intent, study design, patient safety, or may affect a participants willingness to continue participation in the study is considered an amendment, and therefore will be written and filed as an amendment to this protocol and/or informed consent form. All such amendments will be submitted to the HREC, for approval prior to becoming effective.

#### 14.5 *Protocol deviations*

All protocol deviations must be recorded in the patient medical record and on the CRF and must be reported to the Principal Investigator. Protocol deviations will be assessed for significance by the Principal Investigator. Those deviations deemed to have a potential impact on the integrity of the study results, patient safety or the ethical acceptability of the trial will be reported to the HREC. Where deviations to the protocol identify issues for protocol review, the protocol will be amended as per section 11.3.

#### 14.6 *Trial closure*

The study may be terminated prematurely by the Principal Investigator or nominee if:

1. On the advice of the Data Safety Monitoring Committee the number and/or severity of adverse events justify discontinuation of the study.
2. New data become available which raise concern about the safety of the study medications, so that continuation might cause unacceptable risks to subjects.

After such a decision, the Investigator must contact all participants within two weeks, and written notification must be sent to the Ethics Committee.

The Coordinating Centre may terminate the study at a study site/s at any time for any of the following reasons:

1. Failure to enroll participants
2. Major protocol violations
3. Inaccurate or incomplete data
4. Unsafe or unethical practices
5. Safe storage of the study products

In the event an Investigator terminates the study prematurely the Coordinating Centre requires the following:

1. Reasons for termination to be provided in writing.
2. All study supplies, including unused medications and CRFs be returned to the Coordinating Centre.
3. All 'Note for Guidance on Good Clinical Practice' (GCP)[18] documents have been provided to the Coordinating Centre.
4. Investigative site must retain all study documents for at least 21 years after written notification to the Coordinating Centre.

### 15. USE OF DATA AND PUBLICATIONS POLICY

Publication of information and/or data related to this protocol in formats including, but not limited to, conference abstracts, posters or presentations; seminars, journal articles, public reports and internet postings, must be submitted to the BLOOM Trial Management Committee for consideration. Proposals for said activities must be within a reasonable time frame of any due dates. Approval for all said activities must have the written permission of the Chair of the Steering Committee or delegate prior to the event.

### Can Brewer's yeast or beta-gLucan increase mOthers Own Milk supply following preterm birth?

STUDY IDENTIFIER: BLOOM

VERSION: 1.1

DATE: 03/05/2023

PRINCIPAL INVESTIGATOR:
A/Prof Luke Grzeskowiak

SPONSORING INSTITUTION:
South Australian Health and Medical Research Institute
SAHMRI Women and Kids
72 King William Road, North Adelaide SA 5006, AUSTRALIA

Australian and New Zealand Clinical Trials Registry: ACTRN12622000968774

#### PROTOCOL AMENDMENTS

| Version | Date of HREC Approval | Amendments |
|---|---|---|
| V1 | 15/06/2022 | |
| V1.1 | | 1) Amended Principal Site Investigators<br>2) Added Additional Site |

**STATEMENT OF COMPLIANCE**

This document is a protocol for a clinical research study. The study will be conducted in compliance with all stipulations of this protocol, the conditions of ethics committee approval, the NHMRC National Statement on Ethical Conduct in Human Research (2007 – updated May 2015) and the Note for Guidance on Good Clinical Practice (CPMP/ICH-135/95).

I agree that the study will be conducted in accordance with the conditions outlined in the protocol (subject to any amendments). I have read and understood the protocol.

I understand that the information in this protocol is confidential. Publication of information related to this protocol in formats including, but not limited to, conference abstracts, posters or presentations; seminars, journal articles, public reports and internet postings, must be submitted to the Study Steering Committee for consideration. Proposals for said activities must be within a reasonable time frame of any due dates. Approval for all said activities must have the written permission of the Chair of the Steering Committee or their delegate prior to the event.

**Investigator's Name:**

**Investigator's Signature:**

**Date:**

**Study Site:**

**COORDINATING CENTRE:**

Chair Steering Committee: A/Prof Luke GRZESKOWIAK

South Australian Health and Medical Research Institute

**Chair Steering Committee signature:**

**Date:**

#### GLOSSARY OF ABBREVIATIONS

| | |
|---|---|
| AI | Associate Investigator |
| ANZNN | Australian and New Zealand Neonatal Network |
| AE | Adverse Event |
| AR | Adverse Reaction |
| CI | Chief Investigator |
| CRF | Case Report Form |
| EPDS | Edinburgh Postnatal Depression Scale |
| GCP | Good Clinical Practice |
| GMP | Good Manufacturing Practice |
| HREC | Human Research Ethics Committee |
| MPAS | Maternal Postnatal Attachment Scale |
| NHMRC | National Health and Medical Research Council |
| NEC | Necrotizing Enterocolitis |
| NICU | Neonatal Intensive Care Unit |
| PSS:NICU | Parental Stressor Scale: NICU |
| SAE | Serious Adverse Event |
| SAR | Serious Adverse Reaction |
| SAP | Statistical Analysis Plan |
| SOP | Standard Operating Procedure |
| STAI-6 | Spielberger State-Trait Anxiety Inventory |
| TGA | Therapeutic Goods Association |
| UAR | Unexpected Adverse Reaction |

### 1. SYNOPSIS

| | |
|---|---|
| **Title** | Use of Brewer's yeast or beta-glucan for Optimising mothers Own Milk supply following preterm birth |
| **Acronym** | BLOOM |
| **Objectives** | To determine if routine administration of brewer's yeast or beta-glucan improves breast milk production following preterm birth |
| **Design** | Multicentre, double-blind, randomised controlled trial |
| **Population** | Mothers of preterm infants born at less than 34 weeks' gestation who intend to provide breast milk for their infant and recruited less than 72 hours postpartum. |
| **Outcomes** | **Primary:** Daily breast milk volume on Day 7 of intervention<br>**Secondary:** Adverse events, administration of supplemental feeds (donor milk or infant formula), maternal serum prolactin levels, breast milk composition, breastfeeding rates after treatment completion and at infant discharge. |
| **Study Duration** | The study will be completed within a 2-year period. Participants will receive treatment for 7 days. The primary outcome will be evaluated on day 7. Participants and their infants will be followed until the infant reaches term corrected age or is discharged home from the neonatal unit (whichever occurs first). |
| **Interventions** | **Groups:** Brewer's yeast (1680 mg/day)<br>Beta-glucan (250 mg /day)<br>Placebo<br>**Route:** Enteral<br>**Duration:** 7 days |
| **Sample size** | 99 women (33 in each arm) |

### 1. INVESTIGATORS AND FACILITIES

#### 1.1 *Study Investigators*

**A/Prof Luke E GRZESKOWIAK**
Flinders Medical Research Institute
Flinders University of South Australia
Bedford Park SA 5042
AUSTRALIA
&
SA Pharmacy, Flinders Medical Centre
SA Health
Bedford Park SA 5042


*Authorised to sign study protocol and amendments*

**Prof Lisa AMIR**
Judith Lumley Centre
La Trobe University
Bundoora Campus VIC 3086
AUSTRALIA
&
The Royal Women's Hospital
Parkville VIC 3052
AUSTRALIA


*Breastfeeding medical expert for trial*

**A/Prof Alice RUMBOLD**
SAHMRI Women and Kids
Women's and Children's Hospital
North Adelaide SA 5006
AUSTRALIA

**A/Prof Wendy INGMAN**
Adelaide Medical School
University of Adelaide
Woodville SA 5011
AUSTRALIA

**Dr Amy KEIR**
Neonatal Medicine
Women's and Children's Hospital
North Adelaide SA 5006
AUSTRALIA

**Dr Tina BIANCO-MIOTTO**
School of Agriculture Food & Wine
University of Adelaide
Adelaide SA 5005
AUSTRALIA

#### 1.2 *Principal Site Investigators*

**Dr Amy KEIR**
Neonatal Medicine
Women's and Children's Hospital
North Adelaide SA 5006
AUSTRALIA


**Dr Kathryn MARTINELLO**
Department of Neonatology
Flinders Medical Centre
Bedford Park SA 5042
AUSTRALIA


**Prof Lisa AMIR**
Director of the Breastfeeding Service
The Royal Women's Hospital
Parkville VIC 3052
AUSTRALIA


#### 1.3 *Statisticians*

**Dr Emma KNIGHT**
SAHMRI Women and Kids
Women's and Children's Hospital
North Adelaide SA 5006
AUSTRALIA


#### 1.4 *Study Locations*

**Women's and Children's Hospital**
North Adelaide SA 5006
AUSTRALIA

**Flinders Medical Centre**
Bedford Park SA 5042
AUSTRALIA

**The Royal Women's Hospital**
Parkville VIC 3052
AUSTRALIA

### 2. STUDY MANAGEMENT

The South Australian Health and Medical Research Institute is the nominated sponsor for the trial.

The Principal Investigator at each study centre will be responsible for the conduct of the study at their centre including informed consent, recruitment, data collection and maintenance of study documentation. Handling of investigational products will be the responsibility of a clinical trial staff.

The Coordinating Centre Steering Committee will provide direct day-to-day management for the trial. The core Steering Committee will consist of CIs (Grzeskowiak, Amir, Rumbold) and coordinating centre staff including, but not limited to:

- Senior Trials Manager
- Trial Statistician
- Administrator/Data Manager

The core Steering Committee will meet regularly (at least bi-monthly). Every 3-4 months the Clinical Investigators' Group (CIG) will meet. This will compromise all co-applicants and members of the core Steering Committee.

### 3. SERIOUS ADVERSE EVENT AND TRIAL MONITORING COMMITTEES

#### 3.1 *Serious Adverse Event Committee*

A Serious Adverse Event (SAE) Committee blinded to the treatment allocation will be established and chaired by CI Grzeskowiak and will include medical practitioners with expertise in obstetrics and neonatology. The primary role of the SAE Committee is to review all maternal and infant SAEs to determine whether there is any likelihood that involvement in the trial could have contributed. Cause of death will be determined from autopsy results or other hospital summaries by relevant medical personnel. This committee will meet six-monthly (or as required).

#### 3.2 *Data and Safety Monitoring Committee*

An independent Data and Safety Monitoring Committee (DSMC) will be set up to review the yearly progress of the trial and provide feedback to the Steering Committee. The DSMC will review general study progress (recruitment, compliance, loss to follow-up), breast milk supply and key secondary/safety outcomes, and will have access to unblinded data. The DSMC will also provide advice regarding external issues that may impact on the study (for example changes in clinical practice). This committee will meet yearly or as required and will consist of a:

- Neonatologist/Obstetrician
- Clinical Pharmacologist
- Biostatistician

### 4. FUNDING

This study was supported by funding provided by a Channel 7 Children's Research Foundation Fellowship, Australia awarded to LEG (CRF-210323); National Health and Medical Research Council Project Grant, Australia awarded to LEG, WVI, LHA (GNT1165457); as well as a Flinders Innovation Seed Partnership Grant awarded to LEG, LHA, AK, ARR in collaboration between Leiber GmbH, Germany and Flinders University, Australia. Study medications were donated by Leiber GmbH, Bramsche, Germany, who also provided financial support.

### 5. INTRODUCTION AND BACKGROUND

#### *Mothers' own breast milk is best*

Breast milk is considered the optimal nutrition for preterm infants.[1, 2] A compelling body of evidence shows breast milk reduces neonatal morbidity and mortality.[3] Use of mothers' own breast milk during hospitalisation reduces the incidence and severity of preventable morbidities, including necrotizing enterocolitis (NEC), late onset sepsis, chronic lung disease, retinopathy of prematurity, rehospitalisation after discharge, and neurodevelopmental problems in infancy and childhood.[3] For example, a recent Cochrane review showed preterm infants receiving infant formula are three times more likely to develop NEC, which has a mortality rate of 20-40%.[4] Further, the ability for a mother to provide her own breast milk represents an important tangible contribution in a situation where mothers are often unable to provide much infant care in these first few weeks of life. The resultant psychological benefits include greater feelings of attachment, empowerment, and confidence.[5]

#### *Physiology of lactation*

Lactation is regulated by several reproductive (oestrogen, progesterone, placental lactogen, prolactin, and oxytocin) and metabolic hormones (glucocorticoids, insulin, growth, and thyroid). Reproductive hormones act directly on the mammary gland, whereas metabolic hormones have indirect actions through altering endocrine responses and nutrient transfer to the mammary gland.

Development of functional lactation is a multi-stage event. Secretory differentiation (lactogenesis I) occurs in mid- to late pregnancy when differentiation of mammary epithelial cells into lactocytes occurs, conferring the ability to synthesize and secrete key components of human milk. Secretory activation (lactogenesis II) is triggered by the sudden drop in progesterone following delivery of the placenta, accompanied by high levels of circulating prolactin. Milk secretion is copious and occurs between 24–102 hours (average 60 hours) after the birth. This is often described as the milk 'coming in.' Oxytocin is released in response to infant suckling, stimulating contraction of myoepithelial cells around the alveoli and facilitating milk release.[6] Apart from a sudden increase in breast milk volume, the process of secretory activation can be identified by a rapid increase in lactose, citrate, and sodium concentrations in the breast milk. Once lactation is established, an autocrine control (local feedback) mechanism regulates ongoing milk production, meaning supply is largely based on effective removal of milk from the breast.[6]

#### *Preterm birth and insufficient breast milk supply*

Mothers of preterm infants face many challenges in establishing and maintaining an adequate supply of breast milk during their infant's prolonged hospitalisation. This is driven by multiple factors including; physiological immaturity of the breast associated with preterm birth, inability for the preterm infant to breastfeed directly from the breast, and the stress of having an infant admitted to the Neonatal Intensive Care Unit (NICU). Each of these factors has the ability to interfere with establishment of normal milk supply; one previous study demonstrated 82% of women birthing preterm experienced delayed secretory activation.[7] While longer-term breastfeeding outcomes were not collected for this cohort, other studies in mothers of term infants have demonstrated that delayed secretory activation is associated with increased risk of early cessation of breastfeeding.[8]

Some mothers may respond well to non-pharmacological lactation support strategies (i.e. correct use of breast milk pump, increasing expressing frequency) for increasing breast milk supply, but for a large number of mothers, their breast milk supply continues to be insufficient to meet their infant's needs. Insufficient volume of mothers' own breast milk has been partly addressed through the introduction of human milk banks, however there are only five banks across Australia. Further, mothers' own milk has been demonstrated to be superior to donor human milk with respect to composition and bioactivity, highlighting that focusing on supporting mothers to provide their own breast milk to their infants is key to optimising neonatal outcomes.[3]

Given the challenges mothers of preterm infants face with respect to initiation and sustaining lactation, attention has shifted towards the potential role of early non-pharmacological and pharmacological interventions in improving breast milk supply soon after birth. A recent survey of 1876 Australian women found that 60% reported taking galactagogues, known as substances thought to aid in initiating and maintaining adequate milk production, during breastfeeding.[9] This is despite a lack of clear evidence to guide their use.[10-11] The most commonly reported galactagogues in the survey included lactation cookies and brewer's yeast.[9] While lactation cookies may vary in composition, they typically consist of ingredients such as oats, brewer's yeast and flaxseed. The active ingredient of oats is thought to be beta-glucan, which is a glucose polymer present in the cell walls of cereals as well as yeast (i.e. brewers' yeast) and fungi.[12]

While natural galactagogues, such as brewer's yeast, are widely perceived by women to be safer than pharmaceutical galactagogues and are taken by many women,[9] evidence to support their efficacy is largely absent. With respect to brewers' yeast, the likely mechanism of action remains unknown. Some have postulated that it could relate to the high concentration of B vitamins, or the presence of beta-glucan isolated from the cell wall.[13] Studies based on animal models provide evidence that intravenous injection of beta-glucan leads to a significant increase in serum prolactin,[14] providing a potential pathway to changes in breast milk production. The relevance of such findings to oral consumption of beta-glucan in humans, however, remains unclear given the limited bioavailability. In contrast, a number of studies have demonstrated that oral consumption of beta-glucans may have immunomodulatory effects.[12] As immune dysregulation is a common feature of preterm birth[15] and has been associated with impaired lactation in animal models[16], this represents an alternative potential mechanism of action in which brewers' yeast/ beta-glucans could influence breast milk production. Data in lactating women, however, remains scant. A recent clinical trial by Wesolowska et al evaluated the efficacy of a barley malt-based galactagogue (containing a proprietary blend of barley malt and beta-glucan) compared with placebo in mothers of preterm infants.[17] Compared with placebo, those who took the barley malt preparation reported expressing a greater total volume of breastmilk over the 14 day intervention period (6036 ± 498 vs 4209 ± 335; p=0.003). Differences in daily expressed breast milk volume between groups were evident by day 7 of the intervention. No women reported experiencing any side effects during the study. However, the study can only be considered to provide low quality evidence due to a substantial loss to follow-up of 32% across both treatment groups. Therefore, it remains unknown whether natural galactagogues such as brewers' yeast or beta glucan are effective in improving breast milk production following preterm birth.

##### *SUMMARY*

Mothers of preterm infants often struggle in producing enough breast milk to meet the daily feed requirements of their infants, especially in the longer-term. This randomized controlled trial will evaluate the efficacy and safety of two commonly used galactagogues, Brewer's yeast and beta-glucan, compared with placebo in improving maternal breast milk supply following preterm birth.

### 6. STUDY OBJECTIVES

#### 6.1 *Primary objective*

To determine if routine administration of brewer's yeast or beta-glucan improves breast milk production following preterm birth.

### 7. STUDY DESIGN

#### 7.1 *Type of study*

Multicentre, double-blind, randomised controlled trial. Participants, care providers, outcome assessors, trial investigators, and data analysts will be blinded to randomisation group.

#### 7.2 *Number of participants*

The planned sample size is a total of 99 women who gave birth to preterm infants at less than 34 weeks' completed gestation (i.e. 33 per randomised group)

#### 7.3 *Expected duration*

The study will be completed within a 2-year period. Participants will receive treatment for 7 days when the primary outcome will be evaluated. Participants and their infants will be followed until the infant reaches term corrected age or is discharged home from the neonatal unit (whichever occurs first).

##### 7.4 *Primary outcome measures*

Daily breast milk volume on Day 7 following randomisation.

##### 7.5 *Secondary outcome measures*

Adverse events, administration of supplemental feeds (donor milk or infant formula), maternal serum prolactin levels, breast milk composition, breastfeeding rates after treatment completion at infant discharge.

#### 8. STUDY TREATMENTS

##### 8.1 *Treatment arms*

Participants will be randomised to one of three treatment arms, consisting of either brewer's yeast (*Saccharomyces cerevisiae*), beta-glucan (purified from *Saccharomyces cerevisiae*) or placebo (which will be identical to investigational product in appearance).

##### 8.2 *Dosage and route of administration*

All study participants will take three capsules twice daily (six capsules a day) according to the treatment schedule outlined below.

Brewer's Yeast: 1680 mg / day

Beta-glucan: 250 mg / day

Placebo: Microcrystalline cellulose

**Table 1. Dosing regimen**

| **Treatment Arm** | **Morning** | **Night** |
|---|---|---|
| Brewers' Yeast | 3 x 280 mg capsules | 3 x 280 mg capsules |
| Beta-glucan | 2 x placebo capsules<br>1 x 250 mg capsule | 3 x placebo capsules |
| Placebo | 3 capsules | 3 capsules |

To facilitate treatment blinding and aid medication adherence, the medications will be provided in bottles with instructions according to dosing regimen above. All study medications will be identical in appearance.

##### 8.3 *Description*

Study medications will be provided in the form of capsules, identical in appearance.

#### 8.4 *Manufacturer*

Study medications will be manufactured and supplied by Leiber GmbH (Germany).

#### 8.5 *Packaging and labelling*

Participants will be provided sufficient capsules for 7 days of treatment. Each investigational product will be identical in appearance, weight and packaging. The study medications will be packaged and labelled in accordance with GMP including product ID, batch number, expiry date and include the statement “for clinical trial use only”.

#### 8.6 *Medication storage*

Study medications will be stored in a designated secured area at each site and clearly labelled for research purposes only. An inventory will be kept of study medication supplies at all sites.

#### 8.7 *Dispensing and product accountability*

The study coordinator will maintain accurate records of the receipt of all study medication, and when and how much study medication is dispensed and used by each participant in the study. Reasons for departure from the expected dispensing regimen will be recorded.

At the end of the study, there will be final reconciliation of study medications received, dispensed, consumed and returned. Any discrepancies will be investigated, resolved and documented by the study team. Unused study medications will be destroyed in compliance with applicable regulations.

#### 8.8 *Medication adherence*

Medication adherence will be determined informally at the end of the study by the Principal Investigator (PI) of each centre (or nominee), incorporating pill counts. In addition, women will also be provided with a medication diary to keep track of all doses taken (or missed).

#### 8.9 *Concomitant medications/treatment*

Given the absence of any known drug-drug interactions, there are no restrictions on concomitant medications.

### 9. ENROLMENT AND RANDOMISATION

Eligible participants will be provided information on the study by their nurse or midwife who will ask them to complete a Consent to Contact form. Upon providing consent, SAHMRI study staff will contact potential participants to explain the study. The information sheet will describe the purpose of the study, the procedures to be followed, and the risks and benefits of participation. Study staff will conduct the informed consent discussion and will confirm that information provided is understood and answer any questions about the study. Consent will be voluntary and free from coercion. A copy of the consent form will be given to participants and documented in their infant's medical record and study CRF. When all the inclusion and exclusion criteria have been addressed and the eligibility of the participant confirmed, the participant will be randomly assigned to one of three treatment arms.

A record of all mothers screened but not enrolled will be maintained.

#### 9.1 *Inclusion criteria*

Each participant must meet all the following criteria to be enrolled in this study:

- Infant born <34 weeks' gestation (i.e. up to 33+6)
- Intention to provide breast milk
- Between 0 to 72 hours of birth
- Age ≥ 18 years
- Willing and able to comply with all study requirements, including treatment, timing and/or nature of required assessments
- Adequate English language skills
- Signed, written informed consent

#### 9.2 *Exclusion criteria*

- Contraindication to breastfeeding (i.e. HIV)
- Higher order pregnancies (triplet or more)

#### 9.3 *Randomisation procedures*

Women will be allocated to receive either brewer's yeast, beta-glucan, or placebo.

Participants will be randomised using the REDCap Randomisation Module. Participants will be randomised to one of the three treatment arms in a 1:1:1 ratio. The randomisation schedule will be prepared using ralloc.ado in Stata by an independent statistician who is not involved with trial participants or data. The randomisation schedule will be stratified according to study centre.

The schedule will be kept by the independent statistician and the treatment allocation of each randomisation code can be provided to the investigator in case of emergency.

Women must be randomised before starting study treatment. Treatment should be started as soon as possible and preferably within 24 hours of randomisation. Randomisation should occur only after all screening assessments have been performed, participant's eligibility verified and signed consent obtained. Once the randomisation process has been completed as per the instructions in the Study Manual, the woman will be assigned a treatment arm and a study ID.

#### 9.4 *Blinding*

Participants and their care providers, outcome assessors, trial investigators and data analysts will be blinded to randomisation group.

#### 9.5 *Breaking of the study blind*

##### 9.5.1 On study

The randomisation code for an individual participant may only be unblinded in emergency situations, where the Investigator decides a participant cannot be adequately treated without knowing the identity of their treatment allocation. The Principal Investigator must be contacted. All attempts to avoid breaking the code (i.e. withdrawal of treatment) should be made. To break the randomisation code the Investigator must contact the randomisation facility/personnel. The time, date, participant study ID and reason for unblinding must be documented. Events leading to the emergency breaking will be recorded in the serious adverse event (SAE) report form.

##### 9.5.2 Following completion of the study

Trial allocation codes will only be unblinded once all data collected has been entered into the study database for every participant, the database has been finalised and analysis of primary outcome using blinded treatment codes has been completed, except in the case of an emergency, as detailed above.

#### 9.6 *Treatment discontinuation*

Study treatment will be permanently discontinued for any of the following reasons:

- Unacceptable toxicity as determined by the participant or site investigator
- The investigator determined that continuation of treatment is not in the participant's best interest
- Failure to comply with the protocol, the participant declines further study treatment, or withdraws their consent to participate in the study.

#### 9.7 *Participant withdrawal*

Women are free to withdraw themselves and/or their infant from the study at any time. The reasons for withdrawal will be recorded in the CRF and included in the final report. Participants who discontinue treatment or are withdrawn from the study will not be replaced. Whenever possible, permission will be sought from participants who withdraw from the study to obtain as much data for the follow-up period as they will permit.

### 10. STUDY ASSESSMENTS AND PROCEDURES

#### 10.1 *Study procedures*

Participants in the study will be asked to partake in a total of four study related appointments. Appointments will be conducted within the hospital (either within the neonatal unit, postnatal ward, or designated clinical research area), or by telephone (where appropriate).

##### 10.1.1 Demographic/Lifestyle Questionnaire

Background information will be collected in order to describe the characteristics of the study cohort sample. Baseline maternal demographic and lifestyle characteristics include age, height, pre-pregnancy weight, pregnancy weight gain, smoking status, alcohol intake, pre-existing medical conditions (e.g. polycystic ovarian syndrome, diabetes, thyroid disorders), pregnancy complications (e.g. gestational diabetes) and medication use.

##### 10.1.2 Breast Milk Diary

Participants will be asked to maintain a daily breast milk diary throughout each day of the study. This diary will record the number of times each breast was expressed, the method used to express, the volume of expressed milk and if applicable information pertaining to direct breastfeeding. Women will be asked to continue using the diary for 21 days. If the infant directly breastfeeds on day 7 of the study, then test-weighing (i.e. infant weight before and following breastfeed) will be used to estimate breast milk intake.

##### 10.1.3 Breast Health and Milk Expression Questionnaire

Participants will be asked questions relating to methods of breast expression from birth to study enrolment as well as perceived onset of secretory activation. During the study, women will be asked to report data on general breast health (e.g. covers symptoms associated with mastitis), including the ‘Breast milk expression experience measure’ questionnaire.

##### 10.1.4 Postnatal Health Questionnaire

Participants will be asked to detail postnatal health issues during the study (e.g. infections, cold/flu), as well as any medications (prescription or non-prescription), herbal supplements or multivitamins taken.

##### 10.1.5 Mental Health and Wellbeing Questionnaires

Maternal depressive and anxiety symptoms will be collected using standardised validated questionnaires (Edinburgh Postnatal Depression Scale [EPDS], Spielberger State-Trait Anxiety Inventory [STAI-6]). Stress related to infant hospitalisation will be assessed using the Parental Stressor Scale: NICU (PSS:NICU). Maternal-infant attachment will be assessed using the Maternal Postnatal Attachment Scale (MPAS).

Responses to the EPDS will be managed according to our Standard Operating Procedure. In brief, women identified as being at high risk of depression based on responses to the EPDS (i.e. a score of ≥ 13 or positive response to Q10 regarding self-harm) will be notified by study staff of the result, offered written information regarding access to mental health support services and asked to provide consent to share the findings with a trusted healthcare professional in order to obtain further help.

##### 10.1.6 Infant Feeding Practices Questionnaire

Women will complete an infant-feeding questionnaire to determine whether they are breastfeeding exclusively, using a combination of breast- and formula feeding, or formula feeding only.

##### 10.1.7 Medical Record Audit

Maternal and infant case notes will be reviewed to collect data on pregnancy history (e.g. gestational diabetes) and labour/delivery outcomes (e.g. method of delivery, receipt of antenatal steroids prior to delivery, receipt of magnesium sulphate prior to delivery). Infant details collected from medical records include birth weight, gestational age, birth length, head circumference, 1- and 5-minute APGAR scores.

##### 10.1.8 Adverse Events

Information pertaining to potential adverse events or serious adverse events will be collected at each study visit until seven days post-trial completion. Women will also be encouraged to write down any adverse events experienced during the trial in their breast milk production diary.

##### 10.1.9 Medication Compliance Check

Women will be supplied a dosing schedule to record each dose that is taken throughout the study. Further, at the end of each week women will be asked whether they remembered to take all their study medications to evaluate compliance. On day 7 of the study, women will be asked to bring in the study medication packaging, with any remaining capsules to be counted.

##### 10.1.10 Maternal Anthropometrics

Maternal weight, waist circumference, hip circumference, and mid upper-arm circumference will be assessed to evaluate changes in body composition related to the study intervention.

##### 10.1.11 Random Blood Sample

A non-fasted blood sample will be collected to evaluate changes in prolactin, as well as cardiometabolic and inflammatory biomarkers. Blood samples will also be used to obtain maternal DNA. This will be used to perform genetic and epigenetic tests to identify differences in gene variants, DNA methylation and telomere length associated with breast milk supply and galactagogue treatment.

##### 10.1.12 Breast Milk Sample

Breast milk samples (8 mL) will be collected to assess changes in macronutrient composition. A sample will be taken from breast milk expressed between an agreed specified time (9am-11am) in accordance with our Standard Operating Procedure.

###### 10.1.13 Maternal Urine Sample

A urine sample will be collected to assess urinary metabolites and environmental toxins such as phthalates. The sample will be collected, processed and stored in accordance with our SOP.

###### 10.1.14 Maternal Stool Sample

A stool sample will be collected to assess the maternal microbiome in relation to the breast milk and infant microbiomes. The sample will be collected, processed and stored in accordance with our SOP.

###### 10.1.15 Maternal Buccal Swab

A buccal swab will be collected to investigate differences in DNA methylation according to breast milk supply and treatment response to brewer's yeast or beta-glucan. The sample will be collected, processed and stored in accordance with our SOP.

###### 10.1.16 Infant Growth and Anthropometric Measures

Infant head circumference, length and weight measured weekly by medical staff during the infants' hospitalisation will be collected from the infant medical record.

###### 10.1.17 Infant Inpatient Feeding Record (from medical record audit)

The number of days taken to reach full enteral feeds (enteral intake ≥120 mL/kg/day for 3 consecutive days), day of age feeds commenced, days on parenteral nutrition, days on intravenous lipids and type of lipids, type of milk at feed commencement, and at discharge home will be collected from the infant medical record.

###### 10.1.18 Infant Morbidity/Mortality* (from medical record audit)

The number of days in hospital (to first discharge home), postnatal steroid use, grade of intraventricular haemorrhage (IVH), confirmed sepsis, confirmed necrotizing enterocolitis (NEC), grade of retinopathy of prematurity (ROP), surgical procedures and death during first hospitalisation. This data will be collected from the infant medical record.

*All infant clinical data will be collected in accordance with the definitions of the Australian and New Zealand Neonatal Network (ANZNN).[22]

#### 10.2 *Study Assessments*

##### 10.2.1 Study Visit 1 – Randomisation Appointment (Day 0)

Maternal assessments

- Assess study eligibility
- Randomise to treatment group
- Commence Daily Milk Production Diary (10.1.2)
- Medical Record Audit – Pregnancy History (10.1.7)
- Demographic/Lifestyle Questionnaire (10.1.1)

##### 10.2.2 Study Visit 2 (Day 7)

*Protocol window – day 7-9*

Maternal assessments

- Breast Milk Production Diary Check (10.1.2)
- Mental Health and Wellbeing Questionnaires (10.1.5)
    - EPDS; STAI-6; PSS-NICU
- Breast Health Questionnaires (10.1.3)
- Adverse Event Assessment (10.1.8)
- Medication Compliance Check (10.1.9)
- Breast Milk Sample (10.1.12)
- Maternal Urine Sample (10.1.13)
- Maternal Stool Sample (10.1.14)
- Maternal Buccal Swab (10.1.15)
- Maternal Blood Sample (10.1.11)

##### 10.2.3 Study Visit 3 (Day 21 postpartum)

*Protocol window – day 21-23*

Maternal assessments

- Breast Milk Production Diary Check (10.1.2)
- Breast Health Questionnaires (10.1.3)
- Adverse Event Assessment (10.1.8)
- Postnatal Health Questionnaire (10.1.4)

##### 10.2.4 Study Visit 4 (Infant at discharge or term corrected age)

*Protocol window – within 3 days of event*

Maternal assessments

- Infant-Feeding Practices Questionnaire (10.1.6)
- Mental Health and Wellbeing Questionnaires (10.1.5)
    - EPDS; STAI-6; PSS-NICU; MPAS

##### 10.2.5 Medical Record Audit (post-discharge)

- Infant Feeding Data (10.1.17)
- Infant Morbidity/Mortality (10.1.18)
- Infant Growth and Anthropometric Measures (10.1.16)

#### 10.3 *Schedule of assessments – maternal*

| | INTERVENTION PHASE | | | FOLLOW-UP PHASE | |
|---|---|---|---|---|---|
| **MATERNAL** | Screening | Baseline | Day 7 | Day 21 Postpartum | Infant Discharge to Home or Term Corrected |
| **General** | | | | | |
| Eligibility assessment | X | | | | |
| Informed consent | X | | | | |
| Randomisation | | X | | | |
| **Questionnaires** | | | | | |
| Demographic/Lifestyle questionnaire | | X | X | | X |
| Breast milk diary (daily breast milk volume) | | X | X | X | |
| Infant-feeding practices questionnaire | | | | | X |
| Mental health and wellbeing questionnaires<br>- EPDS, STAI-6 | | | X | | X |
| - PSS:NICU | | | X | | X |
| - MPAS | | | | | X |
| Breast health questionnaire | | | X | X | X |
| **Physical Assessments** | | | | | |
| Anthropometric measurements | | | X | | |
| **Case Note Audit** | | | | | |
| Pregnancy/Birth history | | X | | | |
| **Biospecimens** | | | | | |
| Collection of untimed blood for research# | | | X | | |
| Collection of breast milk for research# | | | X | | |
| Collection of urine for research# | | | X | | |
| Collection of stool for research# | | | X | | |
| Collection of buccal swab for research# | | | X | | |

*# Biological samples to be collected at selected sites only. Refer to Standard Operating Procedure for collection procedures.*

#### 10.4 *Schedule of assessments – infant*

| | INTERVENTION PHASE | | | FOLLOW-UP PHASE | |
|---|---|---|---|---|---|
| | Screening | Randomisation / Baseline | Day 7 | Day 21 postpartum | Infant Discharge to Home or Term Corrected |
| **Case Note Audit** | | | | | |
| Infant feeding (daily feeding data) | | X------------------- | ------------------- | ------------------- | -------------------\| |
| Infant morbidity during admission (ANZNN Registry) | | | | | X |
| Anthropometric measurements (as part of routine care) | | X | X | X | X |

### 11. ADVERSE EVENT REPORTING

Any unfavourable and unintended sign, symptom or illness that develops or worsens during the period of the study will be classified as an adverse event (AE), whether it is considered to be related to the study treatment. Adverse events will include unwanted side effects, sensitivity reactions, abnormal laboratory results, injury or inter-current illnesses, and may be expected or unexpected. These will be recorded electronically on the CRF.

Safety evaluations will be conducted each week during the study. Study PI or site-PIs can be directly contacted by the participants if there are any concerns regarding their treatment. The period for adverse event reporting will be from the time of first dose until seven days post final study medication administration. The participants will be followed up face-to-face or by telephone interview at twenty-one days post-partum.

#### 11.1 *Safety reporting for RCT*

Definitions of harm of the EU Directive 2001/20/EC Article 2 based on the principles of ICH GCP apply to this trial.

**Table 2: Adverse Event Definitions**

| | |
|---|---|
| Adverse Event (AE) | Any untoward medical occurrence in a patient or clinical trial participant administered a medicinal product and which does not necessarily have a causal relationship with this product. |
| Adverse Reaction (AR) | Any untoward and unintended response to an investigational medicinal product related to any dose administered |
| Unexpected Adverse Reaction (UAR) | An adverse reaction, the nature or severity of which is not consistent with the applicable product information (eg Investigator's Brochure for an unauthorised product or summary of product characteristics (SPC) for an authorised product. |
| Serious Adverse Event (SAE) or Serious Adverse Reaction (SAR) | Any AE or AR that at any dose:<br>• results in death<br>• is life threatening*<br>• requires hospitalisation or prolongs existing hospitalisation**<br>• results in persistent or significant disability or incapacity<br>• or is another important medical condition*** |
| * The term life threatening here refers to an event in which the patient is at risk of death at the time of the event; it does not refer to an event that might hypothetically cause death if it was more severe (eg a silent myocardial infarction) | |

| |
|---|
| ** Hospitalisation is defined as an in-patient admission, regardless of length of stay, even if the hospitalisation is a precautionary measure for continued observation. Hospitalisation for pre-existing conditions (including elective procedures that have not worsened) do not constitute an SAE<br><br>*** Medical judgement should be exercised in deciding whether an AE or AR is serious in other situations. Important AEs or ARs that may not be immediately life threatening or result in death or hospitalisation, but may seriously jeopardise the participant by requiring intervention to prevent one of the other outcomes listed in the table (eg a secondary malignancy, an allergic bronchospasm requiring intensive emergency treatment, seizures or blood dyscrasias that do not require hospitalisation, or development of drug dependency). |

**Adverse events include:**

- an exacerbation of a pre-existing illness
- an increase in the frequency or intensity of a pre-existing episodic event or condition
- a condition (regardless of whether PRESENT prior to the start of the trial) that is DETECTED after trial drug administration. (This does not include pre-existing conditions recorded as such at baseline – as they are not detected after trial drug administration.)
- continuous persistent disease or a symptom present at baseline that worsens following administration of the trial treatment

**Adverse events do NOT include:**

- Medical or surgical procedures: the condition that leads to the procedure is the adverse event
- Pre-existing disease or a condition present before treatment that does not worsen
- Hospitalisation where no untoward or unintended response has occurred (eg elective cosmetic surgery)
- Overdose of medication without signs or symptoms

#### 11.2 *Seriousness assessment*

When an AE or AR occurs, the investigator responsible for the care of the participant must first assess whether the event is serious. For infant AEs, any deaths will be classified as serious. For maternal AEs, seriousness will be assessed using the definition given in Table 2. If the event is classified as 'serious' then an SAE form must be completed and the Chair of the Steering Committee (or delegated body) notified within one working day.

#### 11.3 *Serious Adverse Events (SAE)*

Determination of the relevant category for reporting a medical event/reaction in the trial will be conducted according to the safety reporting assessment flow chart depicted in

Figure B, 'Safety monitoring and reporting in clinical trials involving therapeutic goods, National Health and Medical Research Council, 2016'. All maternal and infant SAEs are to be reported to the Chair of the Steering Committee, Dr Luke Grzeskowiak within 24 hours of the site becoming aware. Any event that in the opinion of a Principal Investigator may be of immediate or potential concern for a participant's health or well-being will also be reported immediately to the Chair of the Steering Committee. Emergency contacts are listed in section 13.1.

The Serious Adverse Event Committee will review all SAEs. If a mother or infant dies, any post-mortem findings, including histopathology, must be provided to the Coordinating Centre to allow a full independent review.

##### 11.3.1 Severity or grading of adverse events

The severity of all AEs and/or ARs (serious and non-serious) in this trial should be graded using the toxicity grading according to the Common Toxicity Criteria (version 4, 28 May 2009):

1 – Mild
2 – Moderate
3 – Severe
4 – Life threatening
5 – Death

##### 11.3.2 Causality

The SAE Committee must assess the causality of all SAEs in relation to the trial therapy using the definitions in Table 3.

**Table 3: Causality definitions**

| Relationship | Description | Event Type |
|---|---|---|
| Unrelated | There is no evidence of any causal relationship | Unrelated SAE |
| Unlikely to be related | There is little evidence to suggest that there is a causal relationship (eg the event did not occur within a reasonable time after administration of the trial medication). There is another reasonable explanation for the event (eg the participant's clinical condition or other concomitant treatment) | Unrelated SAE |
| Possibly related | There is some evidence to suggest a causal relationship (eg because the event occurs within a reasonable time after administration of the trial medication). However, the | SAR |

| | | |
|---|---|---|
| | influence of other factors may have contributed to the event (eg the participant's clinical condition or other concomitant treatment) | |
| Probably related | There is evidence to suggest a causal relationship and the influence of other factors is unlikely | SAR |
| Definitely related | There is clear evidence to suggest a causal relationship and other possible contributing factors can be ruled out. | SAR |

##### 11.3.3 Expectedness

As it is not expected that the IMP or study protocol will cause any SAEs to mother or infant, all SARs will be classified as unexpected.

#### 11.4 *Emergency contact details*

Dr Luke GRZESKOWIAK – Principal Investigator and Chair of Steering Committee
Flinders Health and Medical Research Institute
Flinders University
Bedford Park SA 5042


Dr Lisa AMIR (Medical Doctor)
Judith Lumley Centre
Level 3, George Singer Building
La Trobe University
Bundoora VIC 3086
AUSTRALIA


#### 11.5 *Australian Therapeutic Goods Association (TGA) and HREC notification*

The Australian TGA requires notification of serious adverse events which are unexpected and deemed to be related to the study treatments. Fatal or life-threatening unexpected adverse events will be notified to the TGA as soon as possible but no later than 7 calendar days after first knowledge by the Coordinating Centre that a case qualifies, followed by an as complete report as possible within 8 additional calendar days. Serious, unexpected adverse events that are not fatal or life-threatening, but deemed related to study treatment shall be filed as soon as possible but no later than 15 calendar days. Serious adverse events that are unrelated to the study treatments shall be included in the end of study report.

The Investigator, or nominee, will also be responsible for reporting any serious adverse events to their Human Research Ethics Committee (HREC) as soon as possible and in any event within 72 hours. In agreeing to the provisions of the protocol, these responsibilities are accepted by the Investigator, or nominee.

If a participant or their infant dies, any post-mortem findings including histopathology, must be provided to the Coordinating Centre.

### 12. STATISTICAL METHODS

#### 12.1 *Sample size estimation*

A sample of 99 women (33 per arm) yields 90% power, 0.025 alpha to show a difference in the mean daily breast milk volume of 150 mL/day (200 mL/day standard deviation) between each of the intervention arms and control, allowing for 10% loss to follow-up. This includes adjustment for a 0.6 correlation between breast milk volume at study entry and breast milk volume on day 7.

#### 12.2 *Statistical Analysis Plan*

A stand-alone Statistical Analysis Plan (SAP) detailing the prespecified analyses to be performed will be produced during the recruitment stage of the study. The SAP will be approved by the Steering Committee prior to any analyses being conducted. Results will be reported according to the CONSORT statement. It is envisaged that the analysis will be undertaken using R and STATA software.

#### 12.3 *Statistical Methods- Outcomes*

The primary analysis will be performed according to the treatment group to which participants were randomised (intention-to-treat principle). A secondary per-protocol analysis will also be performed for each of the primary and secondary outcomes.

The primary outcome of daily breast milk volume on day 7 will be compared between treatment groups using a linear regression. The results will be expressed as a difference in means with a 95% confidence interval and two-sided p-value. Adjustment will be made for baseline breast milk volume and the randomization strata (study centre). A p-value of less than 0.05 will be considered to indicate statistical significance. Analysis of secondary outcomes will use log-binomial regression models for binary outcomes and linear regression models for continuous outcomes with adjustment for stratification variables and other pre-specified prognostic baseline variables. Results will be presented as relative risks and differences in means respectively, along with 95% confidence intervals. Missing data will be addressed using multiple imputation. Sensitivity analyses will also be performed using the original unimputed data.

Planned sub-group analyses of the primary and secondary breastfeeding outcomes include:

(i) Plurality (Singleton vs. twins)
(ii) Parity (Primiparous vs. multiparous)
(iii) Infant gestation at birth ($23^{+0}$–$29^{+6}$ vs. $30^{+0}$–$31^{+6}$ vs $32^{+0}$-$33^{+6}$ weeks' gestation)
(iv) Maternal body mass index (Underweight/Normal weight vs. Overweight/Obese)

Effect modification by each of these factors will be assessed separately by including interaction effects with treatment group in the statistical models.

### 13. DATA MANAGEMENT

#### 13.1 *Data collection*

Data entry and study management will be handled using REDCap. Electronic CRFs will be used, with data collected and stored directly in REDCap. Paper-based CRFs will be available for use where needed.

In order to ensure the accuracy of data collected, representatives from the Coordinating Centre and regulatory authorities will have access to source documents (i.e. mother's or infant's medical records). Confidentiality will be maintained at all times.

#### 13.2 *Data storage*

Paper based CRFs will be stored in a locked office at the study site. Only research staff directly involved in the study will have access to the information.

Electronic forms of data will be collected and stored using REDCap. No confidential data are stored on data entry machines. Each study PI and research staff will be provided with their unique security login. Access to electronic data is granted only to research staff according to specific need. Access is only granted to specific sections of the database(s) and at levels relevant to that person. Only specified staff, using tools within REDCap that track all events, can modify the database.

Transaction logs of the databases to a hard drive on another secure server will be made in accordance with the needs of the project. Web servers and database servers are physically separate.

#### 13.3 *Study record retention*

Original/copies of study documents will be retained at the study site or in archives. Documents will be retained for at least 30 years after study completion in line with the data retention schedules for research involving minors. At the completion of this time documentation will be destroyed using confidential document disposal, by shredding with a commercial grade document shredder. The study electronic data will be stored indefinitely on SAHMRI's secure servers with access only granted to authorised study personnel.

#### 13.4 *Genomic Data Sharing Plan*

It is an international best practice in large-scale genomics research to make deidentified sequencing data publicly available "to facilitate the translation of research results into knowledge, products and procedures that improve human health" (NIH). The Gene Expression Omnibus (GEO) is a public repository hosted by the National Center for Biotechnology Information (NCBI) at the National Institutes of Health (NIH) in the United States. It archives and distributes comprehensive sets of high-throughput functional genomic data submitted by the scientific community. Many scientific journals require proof of GEO accession numbers for study datasets before acceptance of a paper for publication. Some government funding institutions such as the NIH also require investigators to submit a prespecified genomic data sharing plan as a condition of public research funding.

In order to adhere to international best research practice and to ensure eligibility of our study for publication in scientific journals, we plan to archive the fully deidentified sequencing datafiles in GEO (or equivalent database). The sequencing data files prepared for GEO will be free of all identifiers that would permit linkages to individual research participants and variables that could lead to deductive disclosure of the identity of individual subjects, in accordance with the NIH Statement on Sharing Research Data (2003) [See NIH website for further details on data sharing here https://grants.nih.gov/grants/policy/data_sharing/].

### 14. ADMINISTRATIVE ASPECTS

#### 14.1 *Regulatory compliance*

This study will be conducted according to the Note for Guidance on Good Clinical Practice (CPMP/ICH/135/95) annotated with TGA comments (Therapeutic Goods Administration DSEB July 2000) and in compliance with applicable laws and regulations. The study will be performed in accordance with the NHMRC Statement on Ethical Conduct in Research Involving Humans (© Commonwealth of Australia 2007), and the NHMRC Australian Code for the Responsible Conduct of Research (©Australian Government 2007), and the principles laid down by the World Medical Assembly in the Declaration of Helsinki 2008. To this end, no participant will be recruited to the study until all the necessary approvals have been obtained and the participant has provided written informed consent. Further, the investigator shall comply with the protocol, except when a protocol deviation is required to eliminate immediate hazard to a subject. In this circumstance the Principal Investigator and HREC must be advised immediately.

#### 14.2 *Confidentiality*

Participant confidentiality is strictly held in trust by the participating investigators and research staff and their agents. This confidentiality is extended to cover testing of biological samples in addition to the clinical information relating to participants. The study protocol, documentation, data and all other information generated will be held in strict confidence. No information concerning the study or the data will be released to any unauthorised third party, without prior written approval of the coordinating centre. Coordinating Centre and regulatory authorities may inspect all documents and records required to be maintained by the Investigator, including but not limited to, medical records and pharmacy records for participants and their infants in this study subject to individuals having obtained approval/clearance through State/National Governments and HREC as required by local laws. The clinical study site will permit access to such records. Clinical information will not be released without written permission of the subject, except as necessary for monitoring by HREC or regulatory agencies.

#### 14.3 *Independent HREC approval*

This protocol and the informed consent document and any subsequent modifications will be reviewed and approved by the HREC of each study site. A letter of protocol approval by HREC will be obtained prior to the commencement of the study, as well as approval for other study documents subject to HREC review.

#### 14.4 *Modifications of the protocol*

This study will be conducted in compliance with the current version of the protocol. Any change to the protocol document or Informed Consent Form that affects the scientific intent, study design, patient safety, or may affect a participants willingness to continue participation in the study is considered an amendment, and therefore will be written and filed as an amendment to this protocol and/or informed consent form. All such amendments will be submitted to the HREC, for approval prior to becoming effective.

#### 14.5 *Protocol deviations*

All protocol deviations must be recorded in the patient medical record and on the CRF and must be reported to the Principal Investigator. Protocol deviations will be assessed for significance by the Principal Investigator. Those deviations deemed to have a potential impact on the integrity of the study results, patient safety or the ethical acceptability of the trial will be reported to the HREC. Where deviations to the protocol identify issues for protocol review, the protocol will be amended as per section 11.3.

#### 14.6 *Trial closure*

The study may be terminated prematurely by the Principal Investigator or nominee if:

1. On the advice of the Data Safety Monitoring Committee the number and/or severity of adverse events justify discontinuation of the study.
2. New data become available which raise concern about the safety of the study medications, so that continuation might cause unacceptable risks to subjects.

After such a decision, the Investigator must contact all participants within two weeks, and written notification must be sent to the Ethics Committee.

The Coordinating Centre may terminate the study at a study site/s at any time for any of the following reasons:

1. Failure to enroll participants
2. Major protocol violations
3. Inaccurate or incomplete data
4. Unsafe or unethical practices
5. Safe storage of the study products

In the event an Investigator terminates the study prematurely the Coordinating Centre requires the following:

1. Reasons for termination to be provided in writing.
2. All study supplies, including unused medications and CRFs be returned to the Coordinating Centre.
3. All 'Note for Guidance on Good Clinical Practice' (GCP)[18] documents have been provided to the Coordinating Centre.
4. Investigative site must retain all study documents for at least 21 years after written notification to the Coordinating Centre.

### 15. USE OF DATA AND PUBLICATIONS POLICY

Publication of information and/or data related to this protocol in formats including, but not limited to, conference abstracts, posters or presentations; seminars, journal articles, public reports and internet postings, must be submitted to the BLOOM Trial Management Committee for consideration. Proposals for said activities must be within a reasonable time frame of any due dates. Approval for all said activities must have the written permission of the Chair of the Steering Committee or delegate prior to the event.

18. TGA. Note for Guidance on Good Clinical Practice (CPMP/GCP/135/95) annotated with Therapeutic Goods Administration (TGA) comments. In: DSEB, ed2000.

BLOOM

### Can Brewer's yeast or beta-gLucan increase mOthers Own Milk supply following preterm birth?

### Statistical Analysis Plan

| | |
|---|---|
| **Trial registration** | https://www.anzctr.org.au/Trial/Registration/TrialReview.aspx?id=384157 |
| **SAP version** | 1.0, 18/02/2025 |
| **Revision history** | - |
| **Funding** | Channel 7 Children's Research Foundation Fellowship (CRF-210323)<br>Flinders Innovation Seed Partnership Grant, Flinders University<br>Leiber GmbH, Bramsche, Germany |
| **SAP Authors** | Associate Professor Luke Grzeskowiak<br>Chief Investigator<br>College of Medicine and Public Health, Flinders University<br><br>Professor Lisa Amir<br>Chief Investigator<br>Judith Lumley Centre, School of Nursing & Midwifery, La Trobe University<br><br>Associate Professor Lisa Yelland<br>Biostatistician Senior Research Fellow<br>SAHMRI Women and Kids<br>Co-Leader, Biostatistics Unit<br>South Australian Health and Medical Research Institute |

| | Signature | SAP version | Date |
|---|---|---|---|
| Luke Grzeskowiak | [redacted] | 1.0 | 18/02/2025 |
| Lisa Amir | [redacted] | 1.0 | 18/02/2025 |
| Lisa Yelland | [redacted] | 1.0 | 20/02/2025 |

### 1 PREFACE

This Statistical Analysis Plan (SAP) describes the planned analyses and reporting for the Can Brewer's yeast or beta-gLucan increase mOthers Own Milk supply following preterm birth (BLOOM) trial (1). Any deviations from the planned analysis strategy detailed in this SAP will be documented with reasons in a post-analysis version of the SAP. Any post-hoc analyses which are not identified in this SAP but are completed to support the planned analyses will also be identified.

### 2 STUDY METHODS

#### 2.1 Background

Mothers of preterm infants face many challenges in establishing and maintaining an adequate supply of breast milk during their infant's prolonged hospitalisation. Given the challenges mothers of preterm infants face with respect to initiation and sustaining lactation, attention has shifted towards the potential role of early non-pharmacological and pharmacological interventions in improving breast milk supply soon after birth. While natural galactagogues, such as brewer's yeast, are widely perceived by women to be safer than pharmaceutical galactagogues and are taken by many women, evidence to support their efficacy is largely absent. The BLOOM study has been designed to determine the safety and efficacy of brewer's yeast and beta-glucans, derived from Saccharomyces cerevisiae, on breast milk supply in mothers who have had a preterm birth.

#### 2.2 Trial design

This is a multi-centre, parallel, three-arm (1:1:1 allocation), clinician, researcher and participant blinded randomised controlled trial. Mothers of preterm infants born at less than 34 weeks' gestation who intend to provide breast milk for their infant, who are less than 72 hours following birth and able to give informed consent, are randomised to receive either brewer's yeast, beta-glucan or placebo capsules for seven days. The primary outcome is total expressed breast milk volume over a 24-hour period on day 7 of intervention. Participants and their infants will be followed until the infant reaches term corrected age or is discharged home from the neonatal unit (whichever occurs first).

#### 2.3 Randomisation

Participants (women) are randomly assigned to receive intervention of 1680 mg/day Brewers' Yeast (3 x 280 mg capsules in the morning and 3x280 mg capsules at night) or 250 mg /day Beta-glucan (1 x 250 mg capsule + 2 placebo capsules in the morning, 3 placebo capsules at night) or placebo (microcrystalline cellulose, 3 capsules in the morning and 3 capsules at night) using a secure web-based randomisation implement module via REDCap. The randomisation implement service allocates group assignments according to a computer-generated randomisation schedule, prepared by an independent statistician (not involved in trial analysis) using ralloc.ado version 3.7.6 in Stata 16. Randomisation is stratified by location of enrolment (Women's and Children's Hospital, Adelaide (WCH), Flinders Medical Centre, Adelaide (FMC), The Royal Women's Hospital, Melbourne (RWH) using randomly permuted blocks of varying sizes.

#### 2.4 Blinding

Participants, care providers, outcome assessors, research personnel and data analysts are blinded to randomisation group. In the event of a medical emergency where knowledge of the investigational product is critical to a participant's clinical management, the blind for that participant may be broken by the independent statistician who prepared the randomisation schedule via request of the Safety Monitor to the SAHMRI Research Operations Manager. The Principal Investigator shall be notified of the need to approve an unblinding request but would remain blinded to the group allocation. The time, date, participant study ID and reason for unblinding must be documented. Events leading to the emergency breaking will be recorded in the serious adverse event (SAE) report form and reasons for unblinding will be reported as a post-randomisation characteristic (see Section 4.3).

#### 2.5 Sample size

A sample of 99 women (33 per arm) yields 90% power, 0.025 alpha (type I error) to show a difference in the mean daily breast milk volume of 150 mL/day (200 mL/day standard deviation) between each of the intervention arms and control arm, allowing for 10% loss to follow-up. This includes adjustment for a 0.6 correlation between expressed breast milk volume prior to study entry and total breast milk volume on Day 7 at the end of the intervention phase.

#### 2.6 Timing of primary outcome assessment

The primary outcome will be assessed on Day 7 of the intervention.

#### 2.7 Timing of final analysis and unblinding

The database will be locked for analysis once data collection and cleaning are complete and the final version of this SAP has been approved. Following the database lock, unblinded treatment codes will be made available to the trial statistician and analysis of the outcomes listed in this SAP will be performed.

### 3 STATISTICAL PRINCIPLES

#### 3.1 Framework

All comparisons will be undertaken assuming a standard superiority hypothesis testing framework.

#### 3.2 Statistical interim analyses and stopping guidance

No formal interim analyses are planned.

#### 3.3 Confidence intervals and p values

For each outcome variable, a 95% confidence interval will be reported to express uncertainty about the estimated treatment effect, with the effect taken to be statistically significant if the p-value for the two-sided comparative test is <0.025 (to account for two planned pairwise comparisons per outcome). For the three-arm trial, only comparisons with control arm will be performed (i.e. brewer's yeast vs placebo and beta-glucan vs placebo). In describing the effectiveness of the intervention, multiple hypothesis tests will be performed due to multiple secondary outcomes, subgroup analyses and sensitivity analyses for the primary outcome. No multiplicity adjustment will be made for the number of secondary analyses. In the absence of a formal procedure for controlling the type-I error rate, less emphasis will be placed on the results of secondary analyses.

#### 3.4 Adherence and protocol deviations

Each woman is instructed to take three capsules twice daily from enrolment for 7 days (i.e. 1 dose = 3 capsules). Each day women are asked to document if doses were taken or missed. According to these responses, adherence with study supplements during the intervention period will be described for each randomised group as follows:

1. Median and interquartile range (IQR) for the number of study doses taken/missed during the intervention phase.
2. Median and IQR for the percentage of study capsules consumed during the intervention period (100 x number doses taken /number of doses expected to be taken). The number of doses expected to be consumed is defined as the length in days of the intervention period multiplied by two, with women asked to take the study medications twice daily (3 capsules each time). The start of the intervention period is taken to be the date participants were advised to start taking the study medications (this may be the same day as randomisation, or the following day). The end of the intervention period is taken to be Day 7 of the intervention where study medications were continued until this date

3. Frequency and percentage of women with adequate adherence, defined as taking >90% of study medication doses (i.e. at least 13 or more of the assigned doses over the 7 days of intervention)

Frequencies and percentages (of women randomised) will also be presented separately by randomised group for the following protocol deviations:

- Ineligible participant randomised, by reason.
- Randomised in the wrong stratum, by reason.
- Provided the wrong supplements according to randomisation.

#### 3.5 Estimand for the primary outcome

The primary trial objective is to evaluate the efficacy and safety of two commonly used galactagogues, Brewer's yeast and beta-glucan, compared with placebo in improving maternal breast milk supply following preterm birth. The defining attributes of the estimand for addressing this objective are detailed below:

- Target population: Mothers of preterm infants born at less than 34 weeks' gestation who intend to provide breast milk for their infant who are less than 72 hours following birth.
- Treatments: 1680 mg/day Brewers' Yeast or 250 mg/day beta-glucan supplementation.
- Endpoint: Daily breast milk volume on Day 7 following randomisation.
- Population summary: Mean difference (brewers' yeast vs placebo and beta-glucan vs placebo).
- Handling of intercurrent events: Non-compliance with the randomised supplement regime, including treatment discontinuation, taking other supplements, or being administered the wrong study supplements, will be ignored under a treatment policy strategy (consistent with the intention to treat approach detailed in the BLOOM trial protocol).

The secondary per-protocol analysis that was initially planned (1) will not be performed, as such analyses do not align with the estimand framework. Estimands for secondary outcomes are detailed in Section 5.2.

#### 3.6 Analysis population

For each trial outcome the analysis population (or analysis set) indicates the participants who will contribute to the estimation of the population summary measure, as described by the corresponding estimand. Participants will be excluded from the analysis population if the strategy for handling intercurrent events dictates their exclusion (see Section 3.5 for the primary outcome and Section 5.2 for secondary outcome estimand definitions). Participants found to be ineligible after randomisation will remain in the analysis population unless otherwise excluded due to intercurrent events. For the analysis of infant outcomes measured at discharge or term corrected age (see Section 5.1), it will be assumed that infants lost to follow-up or withdrawn from the study who were last known to be alive will survive until this time point, and so will remain in the analysis population.

### 4 TRIAL POPULATION SUMMARIES

#### 4.1 Flow diagram

The following CONSORT flow diagram will be completed to document the flow of participants through the trial.

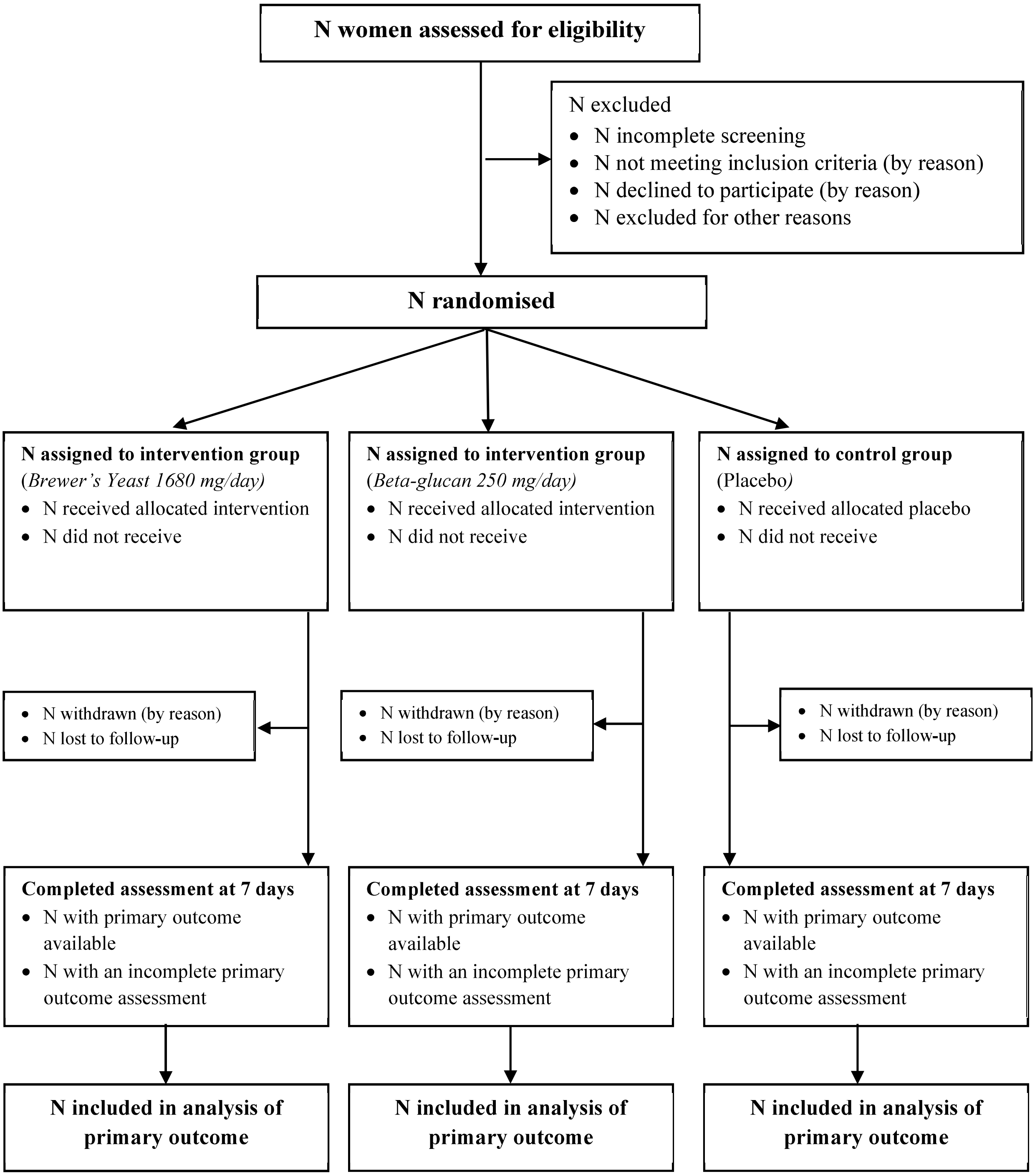

#### 4.2 Baseline characteristics

A descriptive comparison of the randomised groups will be conducted on the baseline characteristics (i.e., characteristics measured at the time of randomisation) presented in Table 1. Comparisons will use all available data, with participant observations attributed to their randomised group irrespective of the occurrence of protocol deviations (see Section 3.4) or intercurrent events (see Section 3.5).

**Table 1.** Baseline characteristics

| **Characteristic** | **Categories** |
|---|---|
| **Maternal** | |
| Recruitment site | Women's and Children's Hospital<br>Flinders Medical Centre<br>Royal Women's Hospital |
| Age, years | Continuous |
| Parity | 1, ⩾2 |
| Multiple pregnancy | Yes/No |
| Pre-pregnancy BMI, $kg/m^2$ | Continuous |
| Born in Australia | Yes/No |
| Aboriginal or Torres Strait Islander | Yes/No |
| Completed secondary education | Yes/No |
| Completed further study | Yes/No |
| Annual household income | ⩽ \$50,000<br>\$50,001 to \$100,000<br>\$100,001 to \$150,000<br>> \$150,001<br>Undisclosed |
| Current smoker | Yes/No |
| Conceived using fertility treatment | Yes/No |
| Pregnancy Complications | |
| Pre-pregnancy diabetes | Yes/No |
| Gestational diabetes | Yes/No/Not applicable |
| Hypertensive disorders of pregnancy | Yes/No |
| Intrauterine growth restriction | Yes/No |
| Gestation at birth, week | Continuous |
| Mode of birth | Vaginal<br>Elective Caesarean Section<br>Emergency Caesarean Section |

| | |
|---|---|
| Hours since birth at time of randomisation | Continuous (hours) |
| Time to first breast milk expression following birth | Continuous (hours) |
| Baseline breast milk volume from single expression prior to intervention, mL | Continuous |
| **Infant** | |
| Sex | Female/Male |
| Birth weight, g | Continuous |
| Birth length, cm | Continuous |
| Head circumference, cm | Continuous |

Means and standard deviations, or medians and interquartile ranges will be reported for continuous variables. Frequencies and percentages will be reported for categorical variables. The clinical importance of any observed imbalances will be noted.

#### 4.3 Post-randomisation characteristics

A descriptive comparison of the randomised groups will be conducted on the post randomisation characteristics relating to the 7-day intervention period presented in Table 2. Comparisons will use all available data, with participant observations attributed to their randomised group irrespective of the occurrence of protocol deviations (see Section 3.4) or intercurrent events (see Section 3.5).

**Table 2.** Post-randomisation characteristics during the intervention period

| Characteristic | Variable Type |
|---|---|
| **Maternal** | |
| Any galactagogue medications (prescription or non-prescription) taken | Binary |
| Type of galactagogue medication taken | Binary |
| Average number of breast milk expression episodes per day | Continuous |
| Infant(s) ever put to the breast | Binary |
| Frequency of double breast expression | Categorical (Never, Hardly ever [>0-20% times a day], Sometimes [20-80% times a day], Mostly/Always [80-100% of times a day]) |
| Study adherence (90% or above percentage of dose expected to take during the intervention period) | Binary |
| Type of side effects reported | Binary |

| | |
|---|---|
| **Other** | |
| Unblinded before primary outcome assessment | Binary |
| Which study group believed to be in | Categorical (Intervention/Control/Unsure) |

Means and standard deviations, or medians and interquartile ranges will be reported for continuous variables. Frequencies and percentages will be reported for categorical variables. The clinical importance of any observed imbalances will be noted.

### 5 ANALYSIS

#### 5.1 Outcome variables

The primary and secondary outcomes for the trial are summarised in Table 3.

**Table 3.** Primary and secondary trial outcomes

| # | Outcome | Time-point measured | Variable type |
|---|---|---|---|
| | **Primary** | | |
| 1 | Expressed breast milk volume over a 24 hour period | Day 7 post intervention | Continuous |
| | **Secondary Maternal** | | |
| | **Maternal breastfeeding outcomes** | | |
| 2 | Expressed breast milk volume over a 24 hour period | Day 21 postpartum | Continuous |
| 3 | Continued any breast milk expression and/or direct breastfeeding | Within the 24 hours before infant discharge or term corrected age | Binary |
| | **Safety outcomes** | | |
| 4 | Maternal side effects (any) | Day 7 post intervention | Binary |
| 5 | Maternal serious adverse events | Day 7 post intervention | Binary |
| 6 | Maternal mortality | Day 7 post intervention | Binary |
| 7 | Infant mortality | Infant discharge or term corrected age | Binary |
| | **Infant outcomes** | | |
| 8 | Total cumulative volume of enteral feeds consisting of formula or donor milk since randomisation | Day 1-7 post intervention / day 1 post intervention to day-21 postpartum / Day 1 to Infant discharge or term corrected age | Continuous |

| | | | |
|---|---|---|---|
| 9 | Total cumulative volume of enteral feeds consisting of mothers own breast milk since randomisation | Day 1-7 post intervention / day 1 post intervention to day-21 postpartum / Day 1 to Infant discharge or term corrected age | Continuous |
| 10 | Infant received exclusively mothers own breast milk as enteral feeds since randomisation | Day 7 post intervention / day 21 postpartum / Infant discharge or term corrected age | Binary |
| 11 | Type of milk feeding at discharge or term corrected age | Within the 24 hours prior to infant discharge or term corrected age | Categorical (breast milk only; infant formula only; mix of breast milk and infant formula) |
| 12 | Weight z-score (g)* | Infant discharge or term corrected age | Continuous |
| 13 | Length z-score (cm)* | Infant discharge or term corrected age | Continuous |
| 14 | Head circumference z-score (cm)* | Infant discharge or term corrected age | Continuous |
| 15 | Weight growth velocity (birth to term equivalent) (g/day)** | Infant discharge or term corrected age | Continuous |
| 16 | Length growth velocity (birth to term equivalent) (cm/week)*** | Infant discharge or term corrected age | Continuous |
| 17 | Head circumference growth velocity (birth to term equivalent) (cm/week)*** | Infant discharge or term corrected age | Continuous |
| 18 | Length of stay (number of days) | Infant discharge | Continuous |

**z-score calculated using Fenton TR, Kim JH. A systematic review and meta-analysis to revise the Fenton growth chart for preterm infants. BMC Pediatrics. 2013;13:59.*

*** Calculated as weight at infant discharge or term corrected age (whichever occurs first) – weight at birth, divided by number of days between measurements.*

**** Calculated as length/head circumference at infant discharge or term corrected age (whichever occurs first) - length/head circumference at birth, divided by number of days between measurements, then multiplied by 7.*

#### 5.2 Estimands

The target population and strategy for handling intercurrent events for each outcome estimand vary according to the timing and level of measurement (infant or mother) of the outcome. Trial outcomes can be grouped together as in Table 4 (using outcome reference numbers from Table 3) to simplify the description of these two estimand attributes.

**Table 4.** Target population and intercurrent event handling by outcome variable groupings

| Outcomes | Estimand attributes |
|---|---|
| 1-3: Maternal breast milk outcomes | Target population: Mothers of preterm infants born at less than 34 weeks' gestation who intend to provide breast milk for their infant.<br>Intercurrent events: Non-compliance with the randomised supplement regime will be ignored under a treatment policy strategy. |
| 4-6: Maternal safety outcomes | Target population: Mothers of preterm infants born at less than 34 weeks' gestation who intend to provide breast milk for their infant<br>Intercurrent events: Non-compliance with the randomised supplement regime will be ignored under a treatment policy strategy. |
| 7: Infant safety | Target population: Infants born at less than 34 weeks' gestation<br>Intercurrent events: Non-compliance with the randomised supplement regime will be ignored under a treatment policy strategy. |
| 8-11: Infant feeding outcomes | Target population: Infants born at less than 34 weeks' gestation<br>Intercurrent events: Non-compliance with the randomised supplement regime will be ignored under a treatment policy strategy. |
| 12-18: Infant anthropometric and hospitalisation outcomes | Target population: infants born at less than 34 weeks' gestation who survive to infant discharge or term corrected age.<br>Intercurrent events: Non-compliance with the randomised supplement regime will be ignored under a treatment policy strategy. Postnatal deaths will be excluded from analysis according to the target population attribute. |

For all outcomes, the remaining attributes of the estimand are defined as follows:

- Treatments: Brewer's Yeast 1680 mg/day or Beta-glucan 250 mg/day
- Endpoint: As defined in Table 3.
- Population summary: Mean difference for continuous outcomes and both an adjusted odds ratio and risk difference for binary outcomes (brewers' yeast vs placebo and beta-glucan vs placebo).

#### 5.3 Overall analysis approach

The primary outcome and all continuous secondary outcomes (see Section 5.1) will be analysed using linear regression, with the effect of treatment described as a mean difference with a 95% confidence interval. All binary secondary outcomes will be analysed using log binomial regression, with the effect of treatment described as a relative risk with a 95% confidence interval. For infant outcomes, clustering due to multiple births will be taken into account using generalised estimating equations with an independence working correlation structure. If the number of infants or mothers (as appropriate) experiencing a binary secondary outcome is too small for a regression model to be sensible (less than 5 events in either randomised group) then, regardless of convergence, a Fisher exact test will be performed instead of logistic regression.

#### 5.4 Covariate adjustment

Given recommendations to adjust for variables used to stratify the randomisation when estimating treatment effects (2), analyses will be adjusted for location of enrolment (WCH, FMC, RWH ), treated as a fixed effect in each analysis model. For the primary outcome, adjustment will also be made for expressed breast milk volume from a single expressing episode prior to commencing study medications. For binary outcomes with low to moderate prevalence, it is possible that adjustment for location of enrolment may lead to model non-convergence. In these instances, there will be no adjustment for stratification. Adjusted analyses will not be considered for binary outcomes analysed using a Fisher exact test (see Section 5.3).

#### 5.5 Planned subgroup analyses

For the primary outcome only, analyses will be performed to test for evidence of effect modification by (1) Plurality (Singleton vs. twins), (2) Parity (Primiparous vs. multiparous), (3) Infant gestation at birth ($23^{+0}$–$29^{+6}$ vs $30^{+0}$–$31^{+6}$ vs $32^{+0}$-$33^{+6}$ weeks' gestation), (4) Maternal body mass index (Underweight/Normal weight vs Overweight/Obese). Effect modification will be assessed by including the subgroup variable as well as its interaction with treatment group into the linear regression model for the primary outcome. For each potential effect modifier, the p-value for the interaction term with treatment group will be reported. Independent of the statistical significance of the interaction p-value, estimates of the treatment effect with 95% confidence intervals will be reported for each subgroup. Subgroup analyses that were initially planned for secondary breastfeeding outcomes (1) will not be performed due to the small sample size and lack of power for assessing effect modification.

#### 5.6 Data transformations and outlying values

No data transformations are planned for continuous outcomes. If the outcome does not follow a normal distribution, the sample size should be sufficient for the central limit to apply. If there is evidence of non-constant variance, robust variance estimation will be used. Outliers will be queried before the database lock. Unless confirmed as a data entry error, outliers will not be excluded from any analyses.

#### 5.7 Methods for handling missing data

Missing data will be summarised descriptively by treatment group for all baseline characteristics (Section 4.2), post-randomisation characteristics (Section 4.3), and outcome variables (Section 5.1). For the primary outcome, missing data will be addressed using multiple imputation implemented under a missing at random (MAR) assumption. Imputation will be performed using fully conditional specification to create 100 complete datasets for analysis. Imputation models will be limited by sample size but will include analysis variables and possibly auxiliary variables associated with missing data. A complete case analysis will also be performed for completeness. For secondary outcomes, analyses will be performed on the available data only.

#### 5.8 Analysis Software

All analyses will be performed using Stata 18 or later (College Station, TX: StataCorp LP) and R 4.3.3 or later (R Foundation for Statistical Computing, Vienna, Austria).
