## Supplemental Table S1 for "Effect of brewers’ yeast or beta-glucan derived from *Saccharomyces cerevisiae* on breast milk supply following preterm birth: The BLOOM randomised controlled trial"

**Table S1: BLOOM Study treatment dosing regimen**

| **Treatment Arm** | **Morning** | **Evening** | **Total amount active ingredient**  **per day** |
| --- | --- | --- | --- |
| Brewers’ Yeast | 3 x 280 mg capsules | 3 x 280 mg capsules | 1680 mg |
| Beta-glucan | 1 x 250 mg capsule  2 x placebo capsules | 3 x placebo capsules | 250 mg |
| Placebo  (Microcrystalline cellulose) | 3 x placebo capsules | 3 x placebo capsules | - |
