## Supplemental Table S2 for "Effect of brewers’ yeast or beta-glucan derived from *Saccharomyces cerevisiae* on breast milk supply following preterm birth: The BLOOM randomised controlled trial"

**Table S2: Adherence and protocol deviations**

| **Maternal variable** | **Brewers’ Yeast**  **N=36** | **Beta-glucan**  **N=35** | **Placebo**  **N=34** | **Overall**  **N=105** |
| --- | --- | --- | --- | --- |
| Study adherence (90% or above percentage of dose expected to take during the intervention period) | | | | |
| Yes | 26 (72) | 18 (51) | 16 (47) | 60 (57) |
| No | 8 (22) | 15 (43) | 14 (41) | 37 (35) |
| Missing | 2 (6) | 2 (6) | 4 (12) | 8 (8) |
| Median and interquartile range (IQR) for the number of study doses missed during the intervention phase | 14 (13, 14) | 13 (11, 14) | 13 (12, 14) | 14 (12, 14) |
| Median and IQR for the percentage of study capsules consumed during the intervention period (100 x number doses taken /number of doses expected to be taken) | 100 (92.9, 100) | 92.9 (78.6, 100) | 92.9 (85.7, 100) | 100 (85.7, 100) |

Values are n (%) unless otherwise indicated.
