## Supplemental Table S3 for "Effect of brewers’ yeast or beta-glucan derived from *Saccharomyces cerevisiae* on breast milk supply following preterm birth: The BLOOM randomised controlled trial"

**Table S3: Complete case analysis for primary trial outcome**

| Primary outcome - Expressed breast milk volume over a 24-hour period (mL), measured at day 7 post intervention^‡^ | | | | | | | |
| --- | --- | --- | --- | --- | --- | --- | --- |
| Brewers’ yeast | Beta-glucan | Placebo | Brewers’ yeast vs Placebo | P value | Beta-glucan vs Placebo | P value | *n* |
| 598 (320) | 430 (273) | 474 (361) | 55 (-96 to 207) | 0.47 | -51 (-206 to 104) | 0.52 | 83 |

^†^ Results are expressed as adjusted difference in means (adjusted for expressed milk volume from a single expressing episode prior to commencing the study medications and location of enrolment) with 95% confidence interval and 2-sided p-value.
