## Supplemental Table S4 for "Effect of brewers’ yeast or beta-glucan derived from *Saccharomyces cerevisiae* on breast milk supply following preterm birth: The BLOOM randomised controlled trial"

**Table S4: Evidence of Effect Modification for the Primary Trial Outcome**

| **Effect Modifier** | **Brewers’ yeast vs Placebo** | **Beta-glucan vs Placebo** | **P value for the overall interaction term** | ***n*** |
| --- | --- | --- | --- | --- |
| Plurality | | | | |
| Singleton | 66 (-84, 216) | -0 (-154, 153) | 0.07 | 83 |
| Twin | -78 (-476, 319) | -483 (-882, -83) |  |  |
| Parity | | | | |
| Primiparous | 116 (-101, 333) | -80 (-282, 122) | 0.52 | 83 |
| Multiparous | 13 (-215, 241) | -8 (-259, 242) |  |  |
| Gestational age at birth | | | | |
| 23^+0^–29^+6^ | 115 (-135, 365) | 26 (-198, 250) | 0.49 | 83 |
| 30^+0^–31^+6^ | -133 (-404, 138) | -280 (-588, 28) |  |  |
| 32^+0^-33^+6^ | 122 (-179, 423) | 42 (-286, 371) |  |  |
| Maternal body mass index | | | | |
| Underweight/Normal weight | -120 (-348, 107) | -8 (-259, 243) | 0.04 | 83 |
| Overweight/Obese | 190 (-7, 387) | -61 (-250, 129) |  |  |

Effect modification was assessed by including the effect modifier as well as its interaction with treatment group into the linear regression model for the primary outcome using the available data. Estimates of treatment effects are expressed as adjusted difference in means (adjusted for location of enrolment and a single expressing episode prior to commencing the study medications) with 95% confidence interval (beta-glucan vs placebo and brewers’ yeast vs placebo). For each potential effect modifier, the p-value for the overall interaction term with treatment group is reported.
