## Supplemental Table S5 for "Effect of brewers’ yeast or beta-glucan derived from *Saccharomyces cerevisiae* on breast milk supply following preterm birth: The BLOOM randomised controlled trial"

**Table S5: Secondary trial outcomes**

| Outcome | Brewers’ yeast | Beta-glucan | Placebo | Brewers’ yeast vs Placebo | P value | Beta-glucan vs Placebo | P value | *n* |
| --- | --- | --- | --- | --- | --- | --- | --- | --- |
| **Infant outcomes** | | | | | | | | |
| Total cumulative volume of enteral feeds consisting of formula or donor milk since randomisation (mL)^*^ | | | | | | | | |
| Day 1-7 post intervention | 156 (245) | 185 (290) | 217 (349) | -54 (-189, 80) | 0.43 | -26 (-182, 131) | 0.75 | 117 |
| Day 1 post intervention to day 21 postpartum | 609 (1272) | 811 (1444) | 953 (1679) | -327 (-991, 337) | 0.34 | -126 (-925, 673) | 0.76 | 117 |
| Day 1 to infant discharge/term corrected age | 2394 (4901) | 3934 (5465) | 2721 (4529) | -290 (-2377, 1797) | 0.79 | 1234 (-1165, 3634) | 0.31 | 117 |
| Total cumulative volume of enteral feeds consisting of mother’s own breast milk as enteral feeds since randomisation (mL) ^*^ | | | | | | | | |
| Day 1-7 post intervention | 319 (286) | 185 (162) | 292 (304) | 29 (-104, 163) | 0.67 | -104 (-215, 7) | 0.066 | 117 |
| Day 1 post intervention to day 21 postpartum | 2815 (1668) | 2181 (1501) | 2395 (1783) | 423 (-404, 1250) | 0.32 | -211 (-1015, 593) | 0.61 | 117 |
| Day 1 to infant discharge/term corrected age | 8488 (5514) | 9226 (6404) | 8729 (7125) | -367 (-3349, 2616) | 0.81 | 336 (-2695, 3366) | 0.83 | 117 |
| Infant received exclusively mothers own breast milk as enteral feeds since randomisation^†^ | | | | | | | | |
| Day 1-7 post intervention^‡^ | 13/41 (32%) | 17/39 (44%) | 13/37 (35%) | 0.90 (0.48, 1.69) | 0.75 | 1.24 (0.71, 2.18) | 0.45 | 117 |
| Day 1 post intervention to day 21 postpartum | 11/41 (27%) | 15/39 (38%) | 10/37 (27%) | 0.96 (0.46, 1.99) | 0.91 | 1.40 (0.72, 2.69) | 0.32 | 117 |
| Day 1 to infant discharge/term corrected age | 9/41 (22%) | 11/39 (28%) | 7/37 (19%) | 1.17 (0.48, 2.82) | 0.73 | 1.47 (0.64, 3.40) | 0.37 | 117 |
| Weight z-score (g) at discharge/term corrected age^*, §^ | -0.69 (0.94) | -1.10 (1.34) | -0.83 (1.64) | 0.08 (-0.51, 0.67) | 0.79 | -0.35 (-1.00, 0.31) | 0.30 | 113 |
| Length z-score (cm) at discharge/term corrected age^*, §^ | -0.78 (1.09) | -1.43 (1.82) | -1.08 (1.92) | 0.27 (-0.45, 0.99) | 0.46 | -0.40 (-1.26, 0.45) | 0.35 | 112 |
| Head circumference z-score (cm) at discharge/term corrected age^*, §^ | 0.0 (1.3) | -0.6 (1.6) | -0.2 (1.6) | 0.1 (-0.6, 0.8) | 0.73 | -0.5 (-1.2, 0.2) | 0.19 | 111 |
| Weight growth velocity (birth to term equivalent) (g/day) ^*, ¶^ | 23.4 (6.1) | 22.7 (5.1) | 22.3 (10.0) | 0.8 (-2.9, 4.5) | 0.67 | 0.1 (-3.5, 3.6) | 0.98 | 113 |
| Length to growth velocity (birth to term equivalent) (cm/week) ^*, #^ | 0.7 (0.6) | 0.9 (0.4) | 0.8 (0.6) | -0.1 (-0.4, 0.2) | 0.46 | 0.0 (-0.2, 0.3) | 0.82 | 109 |
| Head circumference growth velocity (birth to term equivalent) (cm/week) ^*, #^ | 0.7 (0.2) | 0.7 (0.2) | 0.7 (0.2) | 0.0 (-0.1, 0.1) | 0.93 | -0.1 (-0.2, 0.1) | 0.28 | 109 |
| Length of stay (number of days) ^*^ | 48.3 (22.5) | 63.3 (31.1) | 54.8 (35.3) | -6.6 (-21.4, 8.2) | 0.38 | 8.3 (-8.2, 25.0) | 0.32 | 113 |
| Length of stay (number of days) ^‖^ | 48.3 (22.5) | 63.3 (31.1) | 54.8 (35.3) | 0.9 (0.7, 1.2) | 0.37 | 1.2 (0.9, 1.5) | 0.33 | 113 |

^*^ Results for continuous outcomes are expressed as mean (SD) by treatment group and adjusted difference in means (adjusted for location of enrolment) with 95% confidence interval and 2-sided p-value. ^†^Results for binary outcomes are expressed as n/N (%) by treatment group and adjusted risk ratios (adjusted for location of enrolment) with 95% confidence interval and 2-sided p-value. ^‡^ Results are unadjusted difference in means since adjustment for location of enrolment led to model non-convergence ^§^ z-score calculated using Fenton and Kim 2013(1)  ^¶^ Calculated as weight at infant discharge or term corrected age (whichever occurs first) – weight at birth, divided by number of days between measurements. ^\#^ Calculated as length/head circumference at infant discharge or term corrected age (whichever occurs first) - length/head circumference at birth, divided by number of days between measurements, then multiplied by 7. ^‖^ Unplanned analysis, using Negative Binomial regression, performed due to highly skewed distribution of outcome. Results are expressed as mean (SD) by treatment group and adjusted incidence rate ratio (adjusted for location of enrolment) with 95% confidence interval and 2-sided p-value.
